# Association analyses of predicted loss-of-function variants prioritized 15 genes as blood pressure regulators

**DOI:** 10.1101/2023.03.13.23287209

**Authors:** Estelle Lecluze, Guillaume Lettre

## Abstract

**Background:** Hypertension, clinically defined by elevated blood pressure (BP), is an important cause of mortality and morbidity worldwide. Many risk factors for hypertension are known, including a positive family history, which suggests that genetics contribute to inter-individual BP variation. Genome-wide association studies (GWAS) have identified >1000 loci associated with BP, yet the identity of the genes responsible for these associations remains largely unknown.

**Methods:** To pinpoint genes that causally impact BP variation in humans, we analyzed predicted loss-of-function (pLoF) variants in the UK Biobank whole-exome sequencing dataset (n=454,709 participants, 6% non-European ancestry). We analyzed genetic associations between systolic or diastolic BP (SBP/DBP) and single pLoF variants (additive and recessive genetic models) as well as with the burden of very rare pLoF variants (minor allele frequency [MAF] <0.01%).

**Results:** Single pLoF variants in ten genes associated with BP (*ANKDD1B*, *ENPEP*, *PNCK, BTN3A2, C1orf145 [OBSCN-AS1]*, *CASP9*, *DBH*, *KIAA1161 [MYORG]*, *OR4X1*, and *TMC3*). We also found a burden of rare pLoF variants in five additional genes associated with BP (*TTN, NOS3, FES, SMAD6, COL21A1*). Except for *PNCK*, which is located on the X-chromosome, these genes map near variants previously associated with BP by GWAS, validating the study of pLoF variants to prioritize causal genes at GWAS loci.

**Conclusions:** Our study highlights 15 genes that likely modulate BP in humans, including five genes that harbor pLoF variants associated with lower BP.

## INTRODUCTION

Hypertension, defined as SBP >130 mmHg and/or DBP >80 mmHg^1^, affects over 1.28 billion people worldwide and is directly or indirectly responsible for ∼13% of all annual deaths^2^.These numbers are alarming, especially because recent predictions suggest that they will continue to rise in the coming years despite simple disease modifying interventions (e.g. diet, physical activity) and many widely available BP-lowering drugs^3^. Regulating BP is clinically challenging because different patients often respond differently to the same treatments. In part, this is explained by the fact that hypertension results from the complex interplay of the cardiovascular, renal, endocrine and neural systems together, but also in response to variable environmental stimuli^4^.

Genetics also contribute to the risk of developing hypertension: a positive family history of hypertension is a predictive risk factor, and both SBP and DBP are heritable traits (*h*^2^ estimates range from 15-40% and 15-30%, respectively)^5^. Over the last 15 years, increasingly large GWAS have identified >1,000 genetic loci associated with BP variation^6–9^. These genetic associations, enriched for non-coding regulatory common variants, often fail to pinpoint the causal DNA sequence variants and genes. However, this limitation of the GWAS approach is not fatal to all downstream analyses. For instance, the development of BP polygenic risk scores (PRS), which in their simplest forms are the weighted sums of BP-associated variants, is agnostic of the underlying biology. PRS can stratify individuals more at risk to develop high BP, and also associate with several co-morbidities such as stroke and myocardial infarction^6^.

For applications such as drug targeting or repurposing, knowing the identity of the candidate causal BP genes at GWAS loci is however essential. One strategy is to painstakingly characterize candidate genes in cellular and animal models using gain- or loss-of-function experimental designs. The emergence of genome editing technologies (e.g. CRISPR/Cas9) has accelerated the pace of these experiments, yet they remain time-consuming. One complementary approach is to identify and characterize DNA sequence variants that are predicted to completely abrogate the functions of a gene. Such predicted loss-of-function (pLoF) variants are instrumental because they yield biological insights and can also help guide the development of new drugs. Indeed, a pLoF variant should mimic the effect of a drug developed to block the same gene, providing an opportunity to study side effects (e.g. pleiotropy) and therapeutic impact.

In this study, we performed genetic association testing between BP variation and pLoF variants identified by whole-exome sequencing (WES) in the UK Biobank. A combination of additive, recessive, and gene-based statistical models implicated 15 genes in the regulation of BP in humans.

## METHODS

### Description of the UK Biobank WES sets

The UK Biobank cohort includes 502,412 individuals from White, Black, Asian, Mixed and other ancestral backgrounds (**Supplementary Table 1**). To perform our analyses, we defined three set of WES data: (i) the initial 200k WES release (N=200,602) was used as a discovery cohort, (ii) WES data from 254,196 individuals newly available in the 500k release were designated as the 300k set and used as a replication cohort, and (iii) the whole 500k set was also used to perform similar analyses. We summarize our study design in **Figure 1**. This project was approved by the Montreal Heart Institute Ethics Committee (protocol #2017-2247). This work was conducted using the UK Biobank resource (Project number 62518).

**Figure 1.**
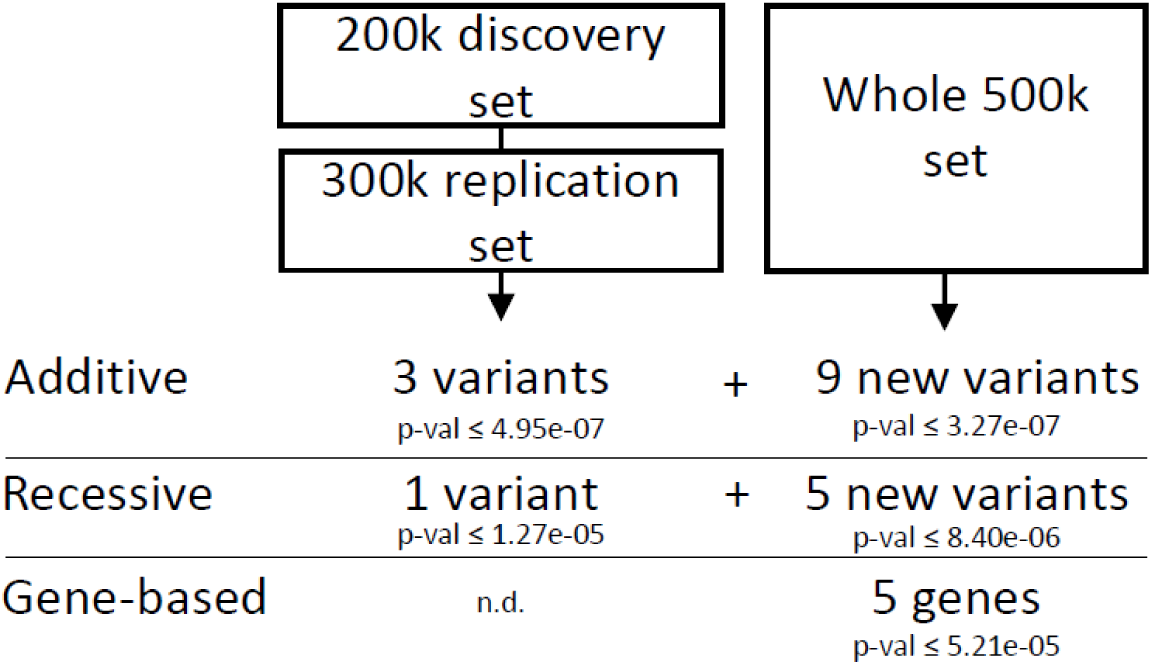
Design of the study to identify predicted loss-of-function (pLoF) variants associated with blood pressure (BP) variation in the UK Biobank. Initially, we performed a discovery analysis in the 200k set followed by a replication analysis in the 300k set. We also analyzed BP associations in the whole 500k set. For each analysis, we report the Bonferroni-corrected statistical threshold. We performed gene-based association tests only in the 500k set. n.d., not done.

### Phenotype normalization

The phenotype was normalized using the whole UK Biobank, including participants that were not included in the WES data. The mean SBP and DBP were calculated using two automated BP measurements (N_SBP_ = 472,371, N_DBP_ = 472,376) or, if unavailable, manual measurements (N_SBP_ = 28,793, N_DBP_ = 28,790). For individuals taking BP medication, 15 mmHg and 10 mmHg were added to their SBP and DBP, respectively. Phenotype normalization was performed separately for each ancestral group. To analyze the SBP and DBP data, a linear regression model was used to calculate residuals of the measure, using sex, age, body mass index, and center of measurement as covariates. A rank-based inverse normal transformation was then applied to these residuals. These inverse normal residuals were scaled to match the standard deviation of the original trait distribution. Therefore, throughout this manuscript, we report genetic effect sizes in standardized mmHg.

### Genotyping array and WES quality control

We then applied quality control measures to the data using plink2, and excluded all unplaced variants, selecting variants with a minor allele count (MAC) >5, filtering out variants and samples with a missing call rate >0.1, and filtering out all variants that had a Hardy-Weinberg equilibrium exact test p-value <1e-15. The snpEff tool was used to identify pLoF variants in the UK Biobank WES dataset. While we tested all pLoF with a MAC >5 in the single variant association tests, we focused our downstream analyses on single pLoF variants with a MAF >0.01%. This effectively excluded two very rare variants in *PYGL* (14:50920952:TCA:T, MAF=3.32e-06 ) and *ZNF804B* (7:89327362:C:T, MAF=4.42e-06)(**Supplementary Table 2**).

### Genetic association (statistical) analyses

Association analyses were performed using the REGENIE (V2.2.4) software^10^. The first step of REGENIE involves fitting a whole-genome regression model to the SBP and DBP data on the complete genotype data. Normalized values of BP were used, and age, sex, and the first 10 principal components were included as covariates. The genotype block was set to 1000 base pairs. The model was used to produce a set of genomic predictions using the additive or recessive models.

The second step of REGENIE involves using WES data to test pLoF variants for association with SBP and DBP, conditional upon the prediction computed in the previous step. The single variant analysis used both an additive and a recessive test, including only pLoF variants with a MAC ≥ 5. Sex, age, and the first 10 principal components were used as covariates, and the genotype data was divided into blocks of 200 base pairs.

For the gene-based analyses, we only included pLoF variants with a MAF ≤ 0.01% and a MAC ≥ 5. We used a burden test to evaluate the aggregated effect of these variants, using the same covariates and genotype block size as in the single variant analyses. pLoF variants within the MHC region were excluded from the final results (hg38: chr6:28,510,120 -33,480,577).

### Databases curation

Each variant or gene of interest were investigated through the curation of GWAS (GWAS catalog^11^), pheWAS (UK Biobank, FINGENN), and aggregated databases (OpenTarget^12^, VannoPortal^13^). We used TOPLD resource^14^ to retrieve linkage disequilibrium (LD) proxies (r^2^≥0.8) of our variants of interest in the European ancestral subset of TOPMed. We also queried single-cell RNA-sequencing data from the Tabula Sapiens Consortium^15^.

## RESULTS

### Single pLoF variants associated with BP in the UK Biobank

We re-analyzed the WES data generated in the UK Biobank (**Figure 1**)^16^. This dataset includes 515,198 pLoF variants found in 454,709 participants, among whom 26,328 participants are of non-European ancestry (**Supplementary Table 1**). Because the WES data was generated by phase, we initiated our discovery effort in the initial 200k set (N=200,602) and replicated the significant results in the 300k set (N=254,196, independent from the 200k set) (**Figure 1** and **Supplementary Table 1**). Under an additive genetic model (**Supplementary Figures 1A and 2A**, **Supplementary Table 2**), this framework identified three replicated pLoF variants in *ANKDD1B*, *ENPEP*, and *PNCK* (**Table 1**). We also tested associations under a recessive genetic model (**Supplementary Figures 1B and 2B**, **Supplementary Table 2**), reasoning that haplosufficient genes would only impact BP when both copies are inactivated by pLoF variants. Since most pLoF variants are rare, we could identify 959 pLoF variants with homozygous carriers, and only the same *ANKDD1B* pLoF variant reached statistical significance in the 200k set and replicated in the 300k set (**Table 1**). To maximize statistical power, we also carried out association testing for the additive and recessive genetic models in the full 500k UK Biobank WES dataset (N=454,709) (**Figure 1**, **Supplementary Figure 3**, and **Supplementary Table 2**). While promising, these results still need to be replicated in external cohorts. The 500k set analyses identified pLoF variants in 10 genes, including seven genes not found in the staged 200k+300k analyses: *BTN3A2, C1orf145 (OBSCN-AS1)*, *CASP9*, *DBH*, *KIAA1161 (MYORG)*, *OR4X1*, and *TMC3* (**Table 1**).

**Table 1.**
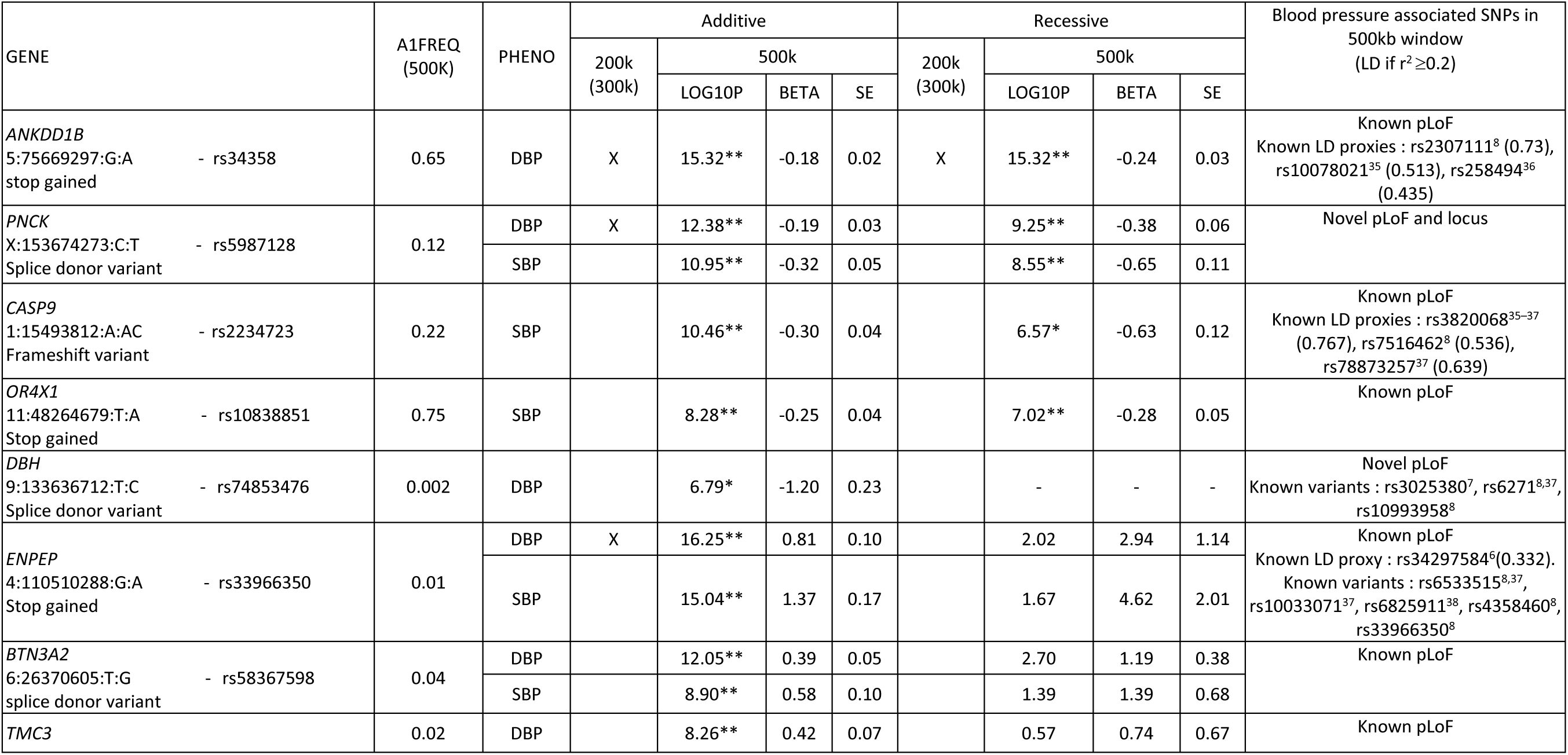
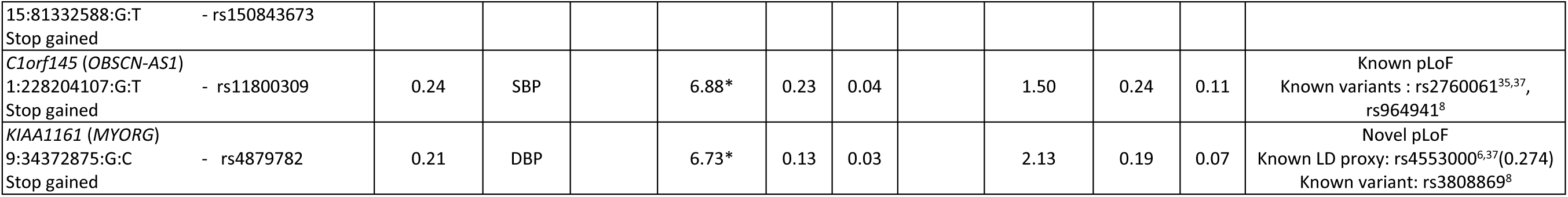
Association results of predicted loss-of-function (pLoF) variants with systolic and diastolic blood pressure (SBP and DBP) (minor allele frequency (MAF) >0.01%). Variants that are significantly associated with a blood pressure trait in the 200k discovery set and replicated in 300k replication set are noted in the “200k (300k)” column. We marked significant P-values after multiple testing correction with a star (additive model : p-value <3.27e-07; recessive model: p-value <8.40e-06), and genome-wide significant variants with two stars (p-value <5e-08). Effect size (BETA) and standard errors are expressed in standardized mmHg. Genomic coordinates (CHR:POS:Allele0:Allele1) are on build GRCh38. The direction of the effect (BETA) is for Allele 1. Linkage disequilibrium (LD) metrics are from European-ancestry individuals in TOPMed.

The association between reduced BP and the *PNCK* splice donor pLoF variant is novel. Although this variant is common (rs5987128, MAF=12%), it might have escaped GWAS detection because it is located on the X-chromosome (which was often excluded from large GWAS consortia efforts). The pLoF variants in *DBH* and *KIAA1161* were not identified in previous GWAS of BP, but other variants located at the same loci have been described (**Tables 1-2**). The seven remaining pLoF variants were reported before (**Tables 1-2**). We confirmed that the BP associations that we identified are driven by the White British participants from the UK Biobank (which represents 94% of the 500k set, **Supplementary Figure 4**) and that their effect sizes are consistent between women and men (**Supplementary Figure 5**).

Half of the pLoF alleles identified in the single-variant analyses are associated with lower BP (**Figure 2**). The strongest effect was observed for carriers of a rare (MAF=0.2%) splice donor pLoF variant in *DBH*, a gene that encodes a dopamine beta-hydroxylase. Rare Mendelian mutations in *DBH* caused orthostatic hypotension 1, and the splice donor pLoF variant identified here (rs74853476, c.339+2T>C) has been characterized as pathogenic in ClinVar^17, 18^. Individuals who carry one copy of this pLoF variant have a DBP that is 1.2 mmHg below the average (**Figure 2**). The four other pLoF variants that decreased BP, as well as the five pLoF that increased BP, are more common (MAF ≥1%) (**Figure 2**). The BP-increasing pLoF variant with the strongest effect – 1.4 mmHg per allele – is a stop gained variant in *ENPEP* (MAF=1%), a gene that encodes an aminopeptidase that can upregulate BP by cleaving angiotensin II.

**Figure 2.**
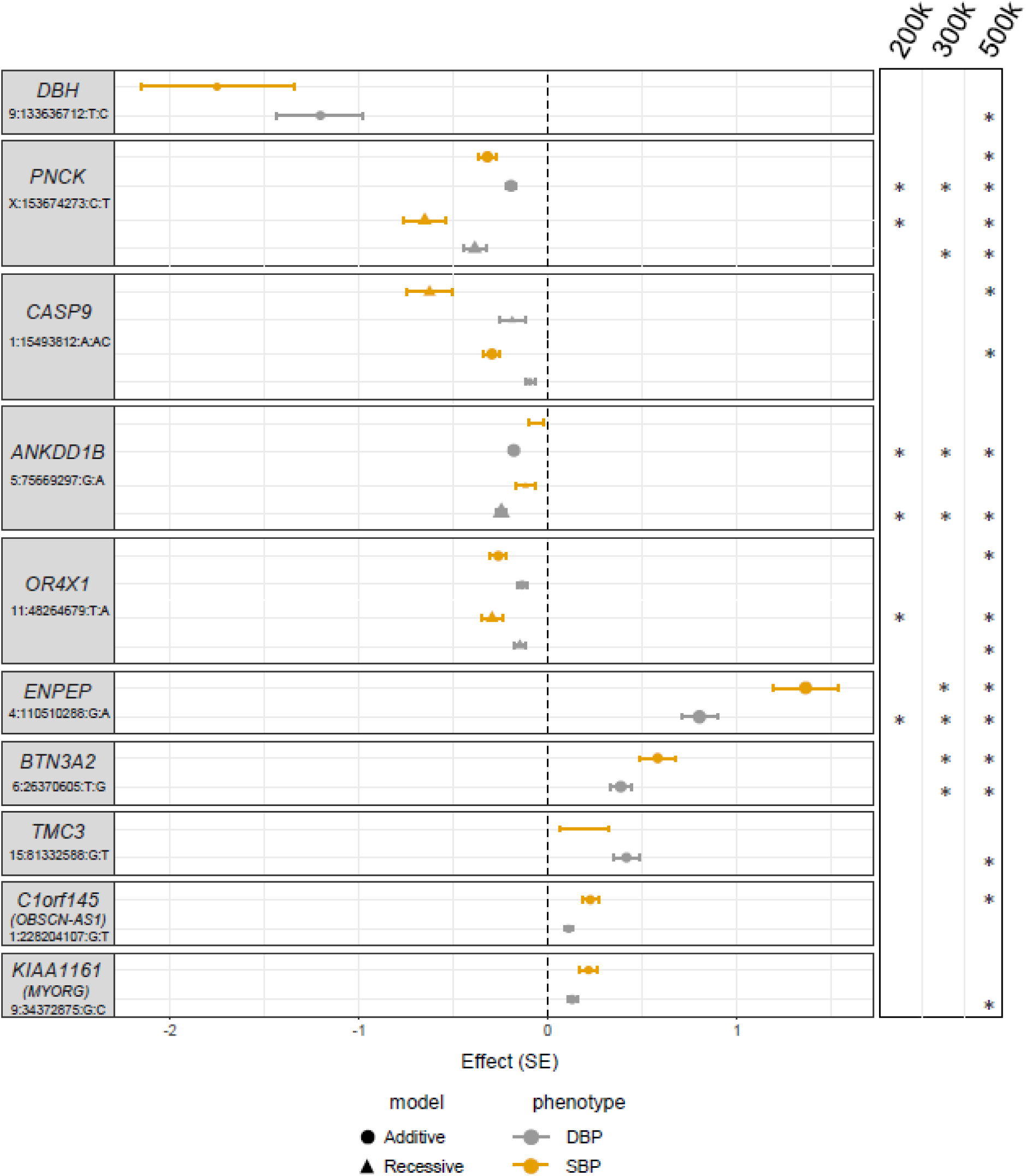
Predicted Ioss-of-function (pLoF) variants associated with blood pressure (BP) variation in single-variant analyses (minor allele frequency (MAF)>0.01%). For each variant, we report the gene, the coordinates on build GRCh38 of the human genome and the corresponding two alleles. Effect sizes and standard errors (SE) are in standardized mmHg. Stars in the panel on the right indicate in which UK Biobank set the variants are significantly associated with BP.

We queried publicly available databases (**Methods**) to determine if the pLoF variants identified in our analyses fell within loci previously associated with other human phenotypes. Except for the *C1orf145 (OBSCN-AS1)* pLoF variant, which is only associated with SBP and hypertension, the remaining variants (or the corresponding genes) are pleiotropic and associate with multiple human traits and diseases, including blood-cell and lipid indices (**Table 2**).

**Table 2.**
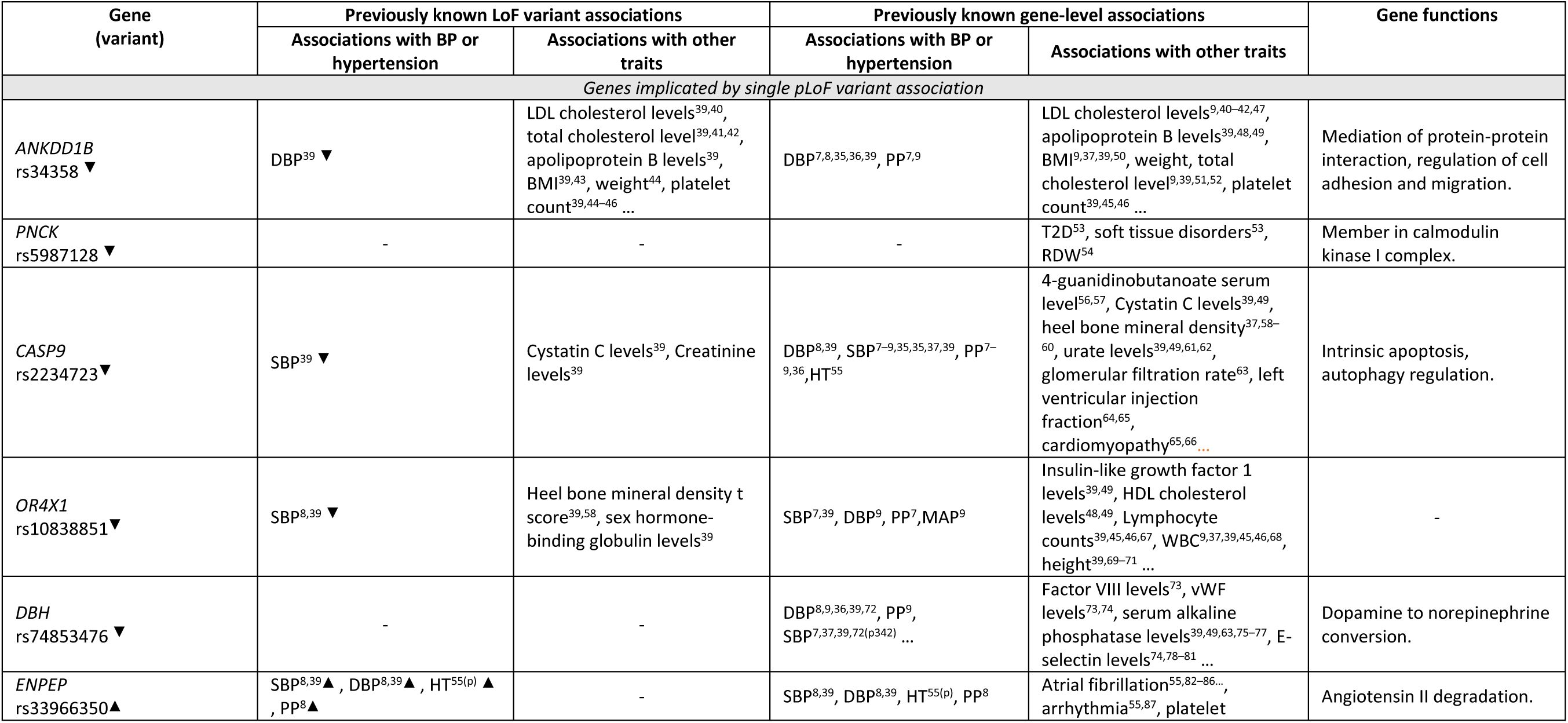
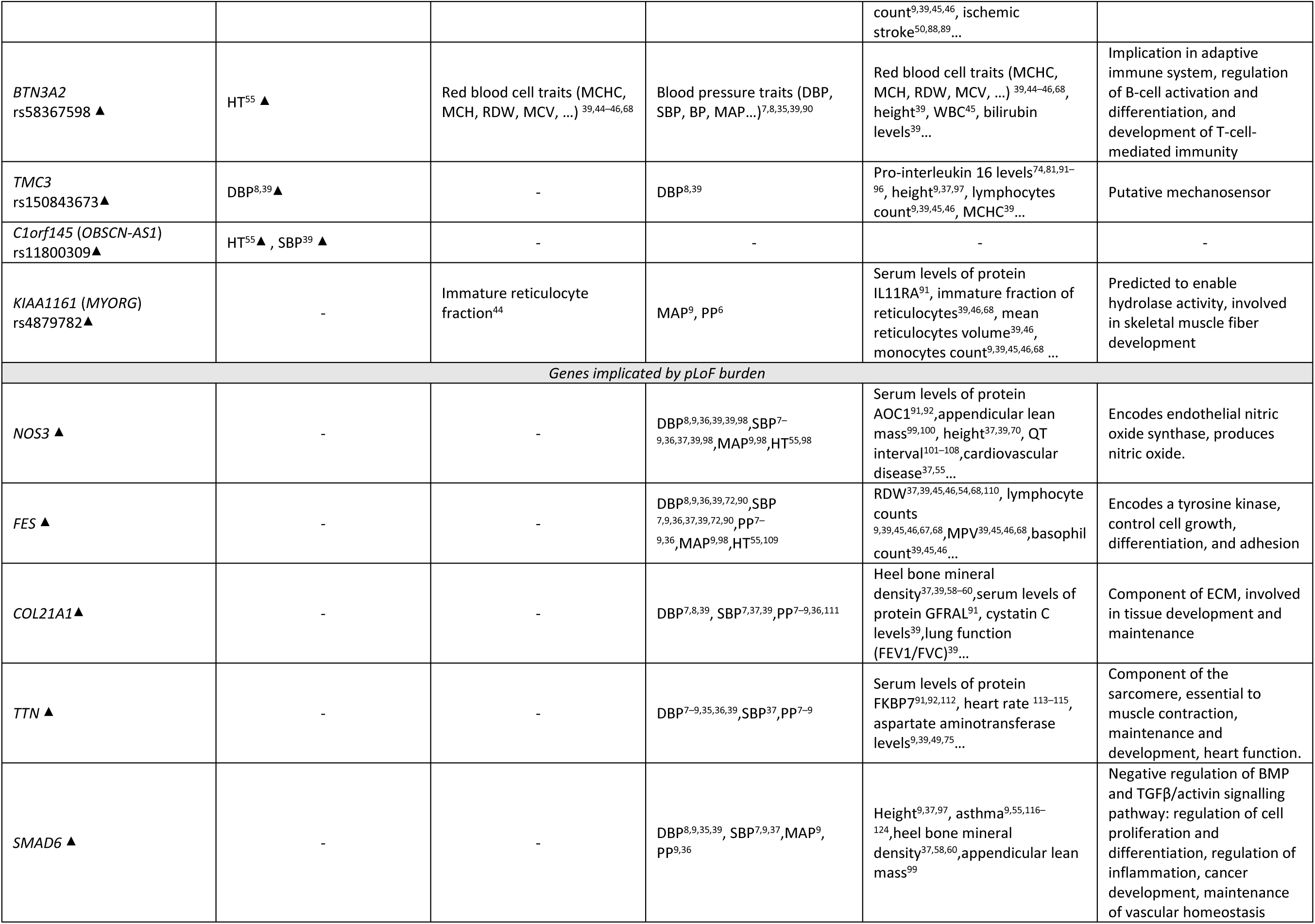
Pleiotropic associations of blood pressure (BP)-associated predicted loss-of-function (pLoF) variants and corresponding genes. This table includes the most significant pleiotropic associations as identified in OpenTargets but is not exhaustive. To populate this table, we queried the Open Targets databases for the pLof variant or its associated gene and included the phenotypes that were associated (pvalue≤10e-8) in the “Previously known LoF variant associations” and “Previously known gene-level associations” columns, respectively. Positive and negative reported effect are represented by and respectively. BMI: body mass index; DBP: diastolic blood pressure; HDL: high density lipoprotein; HT: hypertension; LDL: low density lipoprotein; MAP: mean arterial pressure; MCH: mean corpuscular hemoglobin; MCHC: mean corpuscular hemoglobin concentration; MCV: mean corpuscular volume; PP: pulse pressure; RDW: red blood cell distribution width; SBP: systolic blood pressure; T2D: type 2 diabetes; WBC: white blood cell count.

### Rare pLoF variants associated with BP variation by gene-based analyses

To complement our single-variant association tests and increase power to identify rare pLoF variants associated with BP variation in the 500k set from the UK Biobank, we aggregated pLoF variants from the same gene and performed gene-level association tests (**Figure 1**, **Methods** and **Supplementary Figure 6**). We restricted these analyses to pLoF variants with MAF <0.1% for two reasons: First, including more common variants tended to recover the same variants (and genes) as the single-variant analyses described above. And second, by focusing on rare variants, we limited the possibility to capture association signals arising from residual LD with more common (and likely non-LoF) variants. Using this strategy, we found associations between increased BP traits and the burden of pLoF variants in five genes: *NOS3*, *FES*, *COL21A1*, *SMAD6*, and *TTN* (**Figure 3** and **Supplementary Table 2-3**). Except for *TTN*, for which a burden of pLoF slightly decreases BP, pLoF in the remaining four genes are associated with increase BP (**Figure 3**). Leave-one-variant-out (LOVO) analyses confirmed that these gene-based associations are not due to a single pLoF variant (**Supplementary Figure 7**). *NOS3* represents a strong validation of our strategy as it encodes endothelial nitric oxide (NO) synthase, an enzyme that produces the vasodilator nitric oxide (NO) in the vascular system that helps maintain BP homeostasis. GWAS had previously identified common non-coding variants near these genes, and our gene-based results now nominate *NOS3*, *FES*, *COL21A1*, *SMAD6* and *TTN* as causal regulators of BP in humans (**Table 2**).

**Figure 3.**
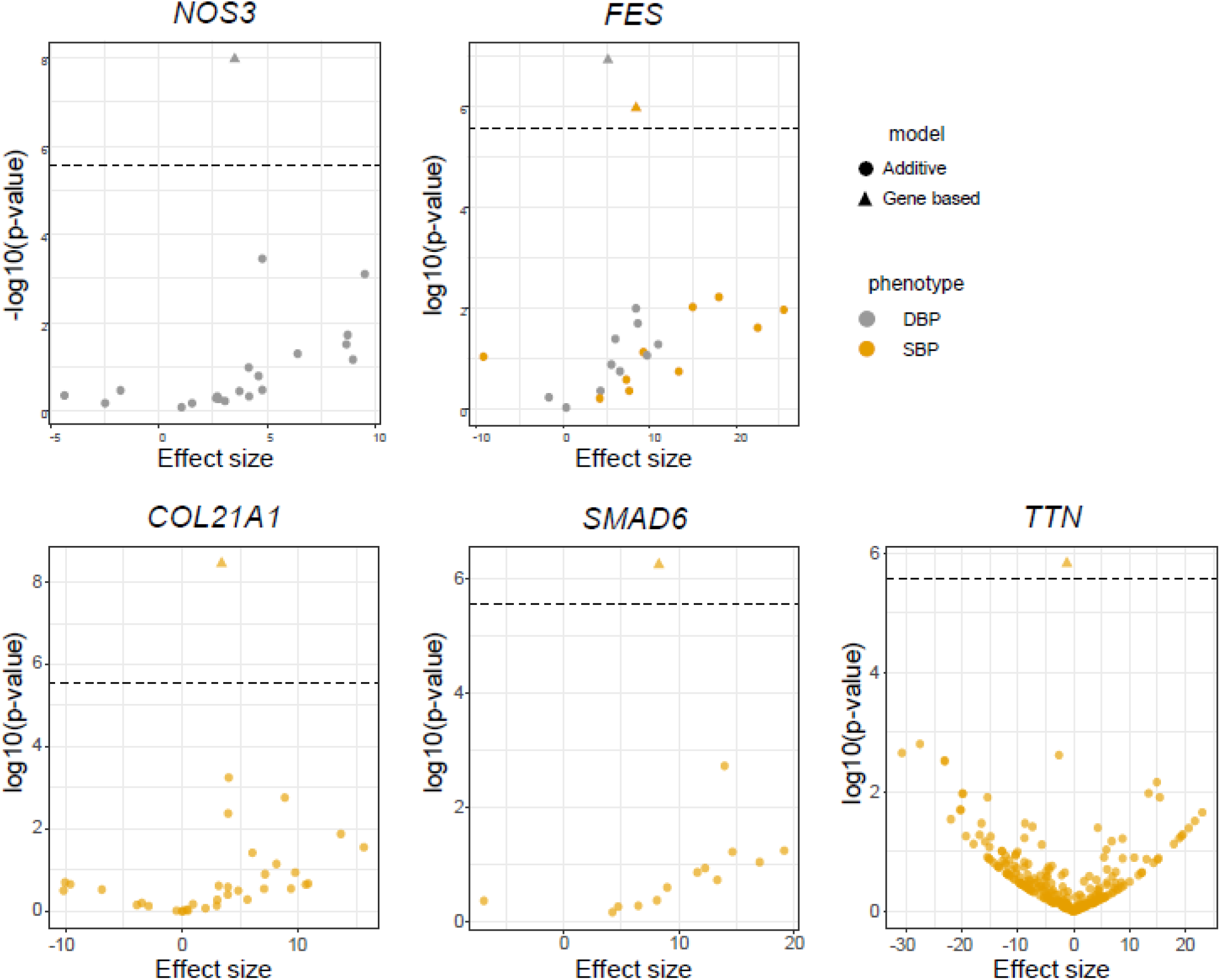
Gene-based association results found five genes that carry rare (minor allele frequency (MAF)≤0.01%) predicted loss-of-function (pLoF) variants associated with blood pressure (BP) variation. On the x-axis, we report the effect size (in standardized mmHg) for the gene-based results as well as for each of the pLoF variant taken individually (additive model). The horizontal dashed line indicates the threshold for gene-based statistical significance (p-val≤5.21e-05).

### Gene expression profiles at single-cell resolution implicate multiple cell-types

To better understand how the 15 genes with pLoF variants (from the single-variant and gene-based tests) may regulate BP, we queried single-cell RNA-sequencing data from the Tabula Sapiens Consortium (**Table 3**)^15^. We found specific expression for *TTN* and *PNCK* in cardiomyocytes, and for *NOS3* and *SMAD6* in endothelial cells, as expected (**Supplementary Figure 8**). The expression of *ENPEP* was largely restricted to kidney epithelial cells, although it was also detectable in smooth muscle cells and pericytes. *BTN3A2* was expressed in T-cells from multiple tissues. The expression patterns of *FES* and *COL21A1* were more diffused: *FES* was expressed in immune (monocytes, macrophages, neutrophils) and endothelial cells, whereas *COL21A1* was found in smooth muscle cells, pericytes and fibroblasts. The expression of the remaining seven genes (*ANKDD1B, CASP9, OR4X1, DBH, TMC3*, *C1orf145 [OBSCN-AS1],* and *KIAA1161 [MYORG]*) was too low in the Tabula Sapiens dataset to be unambiguously assigned to specific cell-types.

**Table 3.** Expression of genes that carry predicted loss-of-function (pLoF) variants associated with blood pressure (BP) in the Tabula Sapiens Consortium single-cell RNA-sequencing dataset. We analyzed the whole dataset (n=483,152 cells, 45 tissues) using cell-type annotations provided by the Consortium. Seven genes could not be unambiguously assigned to specific cell-types: *ANKDD1B, CASP9, OR4X1, DBH, TMC3*, *C1orf145 (OBSCN-AS1),* and *KIAA1161 (MYORG)*. See also **Supplementary Figure 8** for details.

| Gene | Cell-type(s) (Tabula Sapiens) |
| --- | --- |
| *TTN* | Cardiomyocytes |
| *PNCK* | Cardiomyocytes |
| *NOS3* | Endothelial cells |
| *SMAD6* | Endothelial cells (mostly capillary and arterial) |
| *ENPEP* | Mostly kidney epithelial cells. Weaker signal in smooth muscle cells and pericytes. |
| *COL21A1* | Smooth muscle cells, pericytes, fibroblasts. |
| *FES* | Immune cells (monocytes, macrophages, neutrophils), endothelial cells. |
| *BTN3A2* | T-cells |

## DISCUSSION

Hypertension is a central risk factor for stroke, ischemic heart disease, renal dysfunction, dementia and other cardiovascular diseases. Using a combination of different statistical models and the large WES dataset from the UK Biobank, we highlighted 15 genes that likely impact the regulation of BP in humans. Indeed, the list is enriched with genes that encode proteins with well-defined role(s) in BP homeostasis: *NOS3*, *DBH*, and *ENPEP*. These positive controls genes are further discussed in the **Supplementary Notes**, along with *CASP9* (renal functions), genes that have known roles in the cardiovascular system (*SMAD6, COL21A1, TTN*), and genes with more speculative functions in BP regulation and hypertension (*BTN3A2, ANKDD1B, OR4X1, TMC3, C1orf145 [OBSCN-AS1]*, *KIAA1161 [MYORG]*). Below, we discuss two genes of particular interest: *PCNK* and *FES*.

*PNCK* encodes the pregnancy-upregulated non-ubiquitous calcium-calmodulin-dependent kinase (also known as calmodulin kinase 1b [CaMKIB]), a member of the protein serine/threonine kinase family. This gene, a paralog of the canonical *CAMKI* gene, is not well-characterized functionally. Its strongest expression is in the developing brain, but it remains expressed in numerous tissues, including the heart (**Supplementary Figure 8**) and the mammary gland during pregnancy^19^. *PNCK* is also differentially expressed in the placenta of pregnant women with pre-eclampsia, a condition characterized by hypertension^20^. In contrast to *PNCK* and its *CAMKI* family members, genes that encode members of the *CAMKII* family have well-known roles in regulating BP. In vascular smooth muscle cells, CaMKII responds to angiotensin II to increase BP^21^, whereas in endothelial cells, it can phosphorylate eNOS and promote the production of the vasodilator NO^22^. While not specifically developed for PNCK, CaMKI inhibitors have been tested in diabetic and obese mouse models. It would be interesting to know whether the same molecules can interfere with PNCK activity, and whether this affects BP^23^.

*FES* is a proto-oncogene that codes for a protein tyrosine kinase. It is expressed in multiple cell-types, including myeloid and endothelial cells. GWAS have identified SNPs at the *FES* locus that associate with BP and coronary artery disease^9, 24^. *FURIN* is located just upstream of *FES* and encodes a subtilisin-like proprotein convertase with pro-atherogenic functions^25^. Therefore, it represents a more obvious candidate causal gene to explain the GWAS signal at the locus. However, several lines of evidence now suggest that *FES* might also contribute to the GWAS signal for hypertension and CAD risk: (1) statistical and epigenomic fine-mapping has prioritized a SNP (rs12906125) in the promoter of *FES* as the likeliest causal variant at the locus^26^, (2) this variant is a stronger expression quantitative trait locus (eQTL) for *FES* than *FURIN*^27^, (3) CRISPR activation at this variant strongly upregulates *FES* expression and induces endothelial dysfunction^28^, and (4) specific base editing at rs12906125 impacts the expression of *FES*, but not *FURIN*, in endothelial cells treated with TNFα^28^. Our new analyses now show that rare pLoF in *FES*, which are independent from the GWAS signal, are associated with BP, further suggesting that *FES* also contributes to BP regulation and CAD risk. Importantly, our data do not rule out *FURIN* as an equally important local contributor to the GWAS signal. Indeed, while they do not reach statistical significance after multiple testing correction, the 42 pLoF variants in *FURIN* found in the UK Biobank 500k set are nominally associated with BP (burden P-value=0.002).

Many phenotypes, including BP, have already been analyzed in the WES UK Biobank dataset^16, 29^. While there are small differences (e.g. focus on rare pLoF variants, adjustment for BP-lowering drugs, marginal differences in terms of association P-values due to the statistical tests used, no MAF threshold for gene-based tests …), their results are largely consistent with ours. However, because our study is specifically focused on the genetics of BP, we bring into the spotlight 15 candidate BP genes, genes which were not specifically discussed by these other studies since they analyzed 1000s of other phenotypes. We acknowledge several limitations of our study. Most importantly, because 94% of the UK Biobank is of European ancestry, we were not well-powered to identify BP-associated pLoF variants in other ancestral groups. We were also limited in power to test the role of very rare alleles (e.g. singletons and doubletons), and expect that future studies in larger (and more diverse) datasets will make exciting new BP discoveries. Another area for future development is the inclusion of additional classes of genetic variants in pLoF variant association studies. For simplicity, we restricted our analyses to nonsense, frameshift and essential splice site variants, but we recognize that additional variants – such as a subset of missense variants – can also have null function. Including these variants in association tests will broaden the list of genes that we can test for a role in human phenotypic variation.

Motivated by work in animal models and Mendelian genetics, many studies have now considered the impact of pLoF variants on human diseases or traits^30–34^. We extended this framework to the large UK Biobank WES dataset and identified 15 genes likely implicated in BP homeostasis. These genes are expressed in multiple tissues and cell-types, consistent with the network complexity that controls BP. Strikingly, only one of these genes – *PNCK* – is located at a locus that was not previously associated with BP by GWAS (likely because it is on the X-chromosome). This suggests that the true value of pLoF-based studies will not be in identifying new loci, but rather in prioritizing strong candidate genes within GWAS regions, thus streamlining downstream functional experiments. For BP and hypertension more specifically, our data highlight that the putative inactivation of five genes – *ANKDD1B, PNCK, CASP9, OR4X1*, and *DBH* – reduces BP, prompting further analyses to determine if the encoded proteins could be safe and efficacious BP-lowering drug targets.

## Supporting information

Supplemental results

## Data Availability

All data produced are available online at: http://www.mhi-humangenetics.org/en/resources/

## Acknowledgements

We thank the participants from the UK Biobank who generously contributed their data to enable this research. This research was enabled in part by support provided by Calcul Quebec (https://www.calculquebec.ca/en/) and Compute Canada (www.computecanada.ca).

## Supplemental Material

Supplemental Text Supplementary tables

## Declarations

### Ethics approval and consent to participate

This project was approved by the Montreal Heart Institute Ethics Committee (protocol #2017-2247). This research has been conducted using the UK Biobank resource under application number 62518.

### Availability of data and materials

Single-variant and gene-based summary association statistics are available at: http://www.mhi-humangenetics.org/en/resources/

#### Author contributions

Conceptualization: Estelle Lecluze, Guillaume Lettre

Data curation: Estelle Lecluze, Guillaume Lettre

Formal analysis: Estelle Lecluze

Funding acquisition: Guillaume Lettre

Investigation: Estelle Lecluze

Project administration: Guillaume Lettre

Resources: Estelle Lecluze

Supervision: Guillaume Lettre

Validation: Estelle Lecluze

Visualization: Estelle Lecluze

Writing: Estelle Lecluze, Guillaume Lettre

## ABBREVIATIONS

BP: blood pressure
DBP: diastolic blood pressure
GWAS: genome-wide association study
LD: linkage disequilibrium
MAC: minor allele count
MAF: minor allele frequency
mmHg: millimeters of mercury
pLoF: predicted loss-of-function
PRS: polygenic risk score
SBP: systolic blood pressure
WES: whole-exome DNA sequencing

---

**Supplementary Figure 1.**
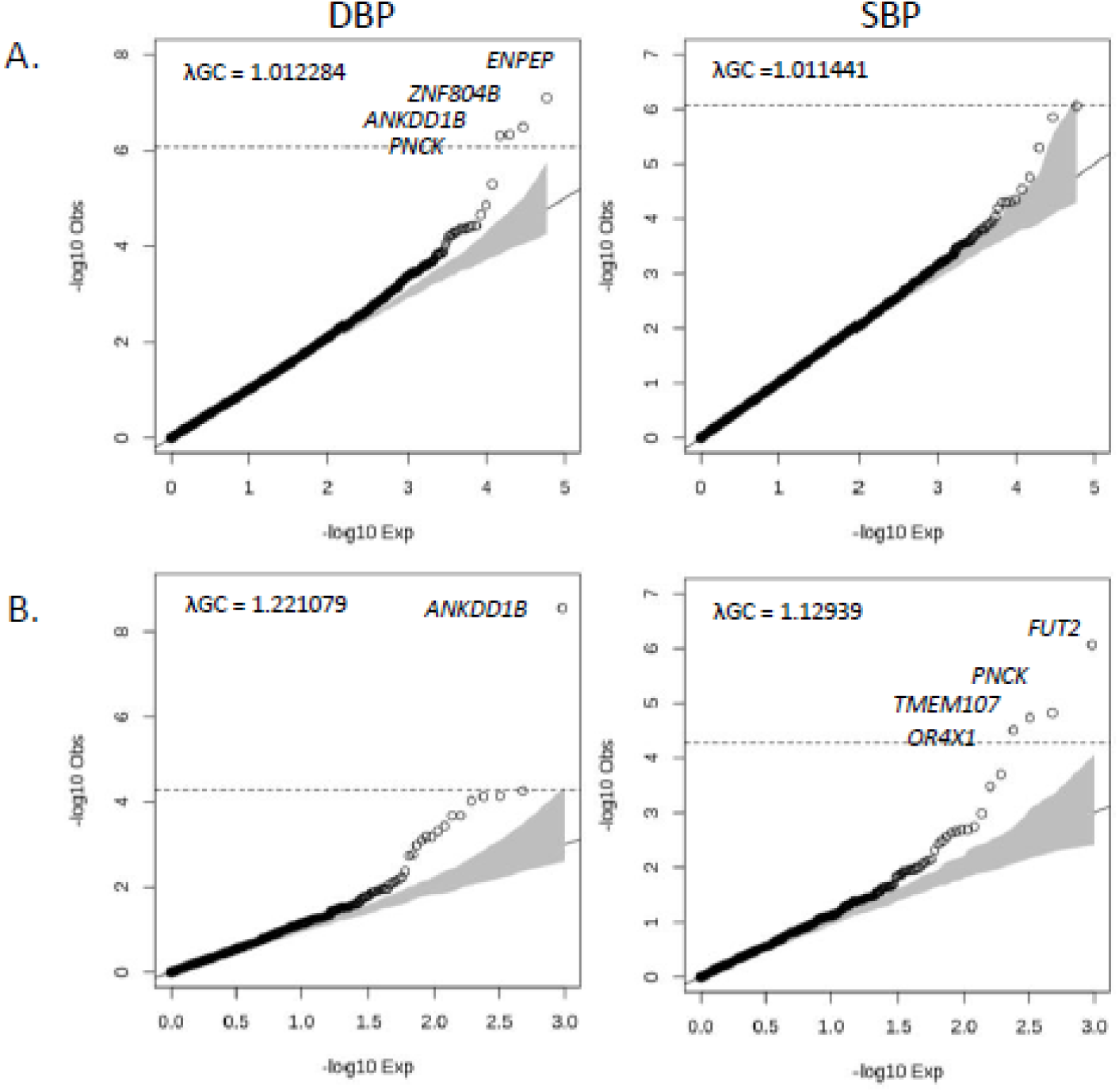
Quantile-quantile (Q-Q) plots for predicted loss-of function (pLoF) variant association results with diastolic blood pressure (DBP) and systolic blood pressure (SBPļ in the 200k (discovery) set (N_sample_=199,558) using **(A)** an additive model (N_variant_=59,418) and **(B)** a recessive model (N_variant_=959). In each plot, the dashed line represents the statistical significance threshold (additive model p-val≤8.41e-07; recessive model p-val≤5.21e-05) and the grey area is the 95% confidence interval.

**Supplementary Figure 2.**
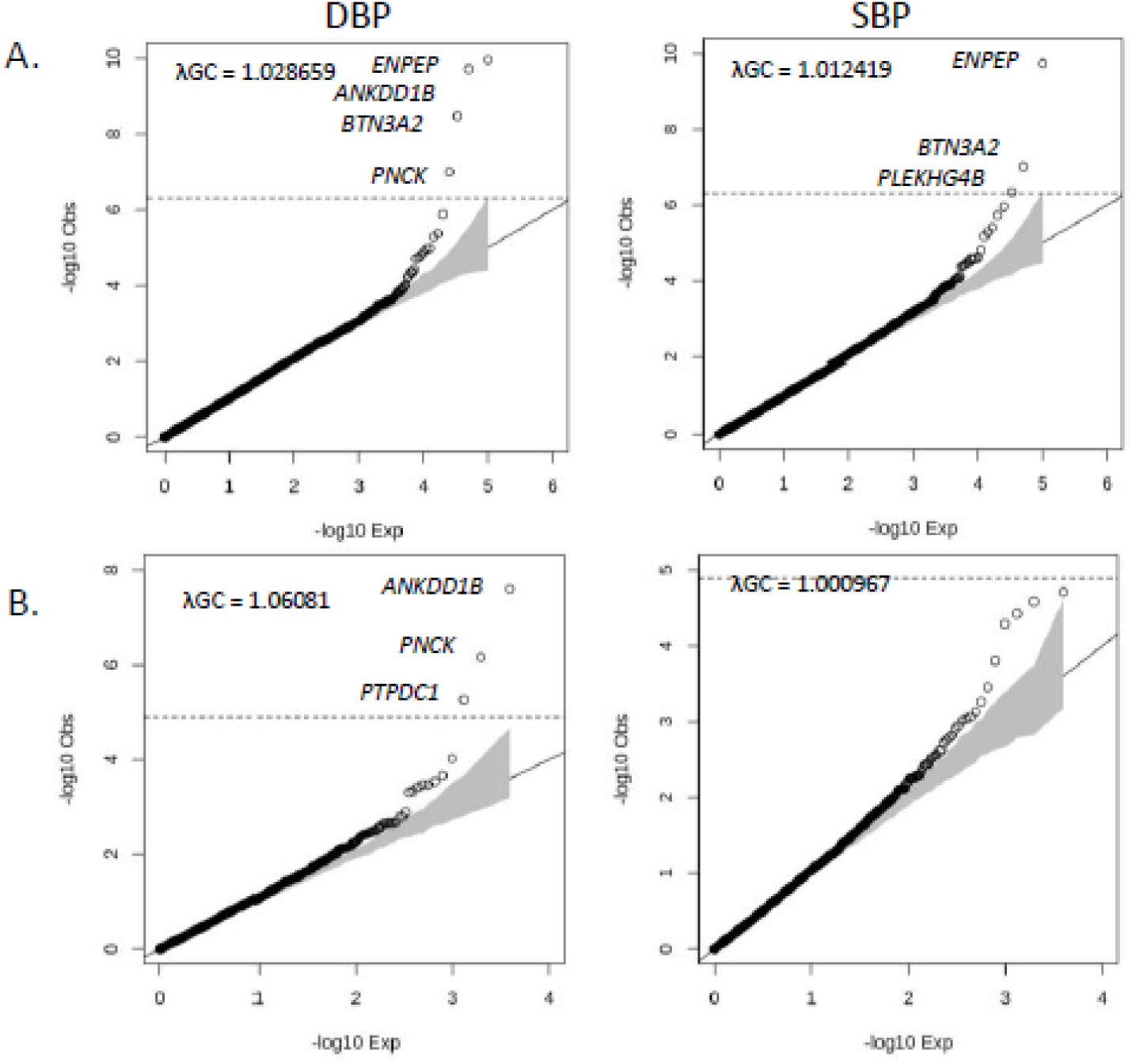
Quantile-quantile (Q-Qļ plots for predicted loss-of function (pLoF) variant association results with diastolic blood pressure (DBP) and systolic blood pressure (SBP) in the 300k (replication) set (N_sample_=252,898) using (A) an additive model (N_variant_=101,030) and (**B**) a recessive model (N_variant_=3,944). In each plot, the dashed line represents the statistical significance threshold (additive model p-val≤4.95e-07; recessive model p-val≤1.27e-05) and the grey area is the 95% confidence interval.

**Supplementary Figure 3.**
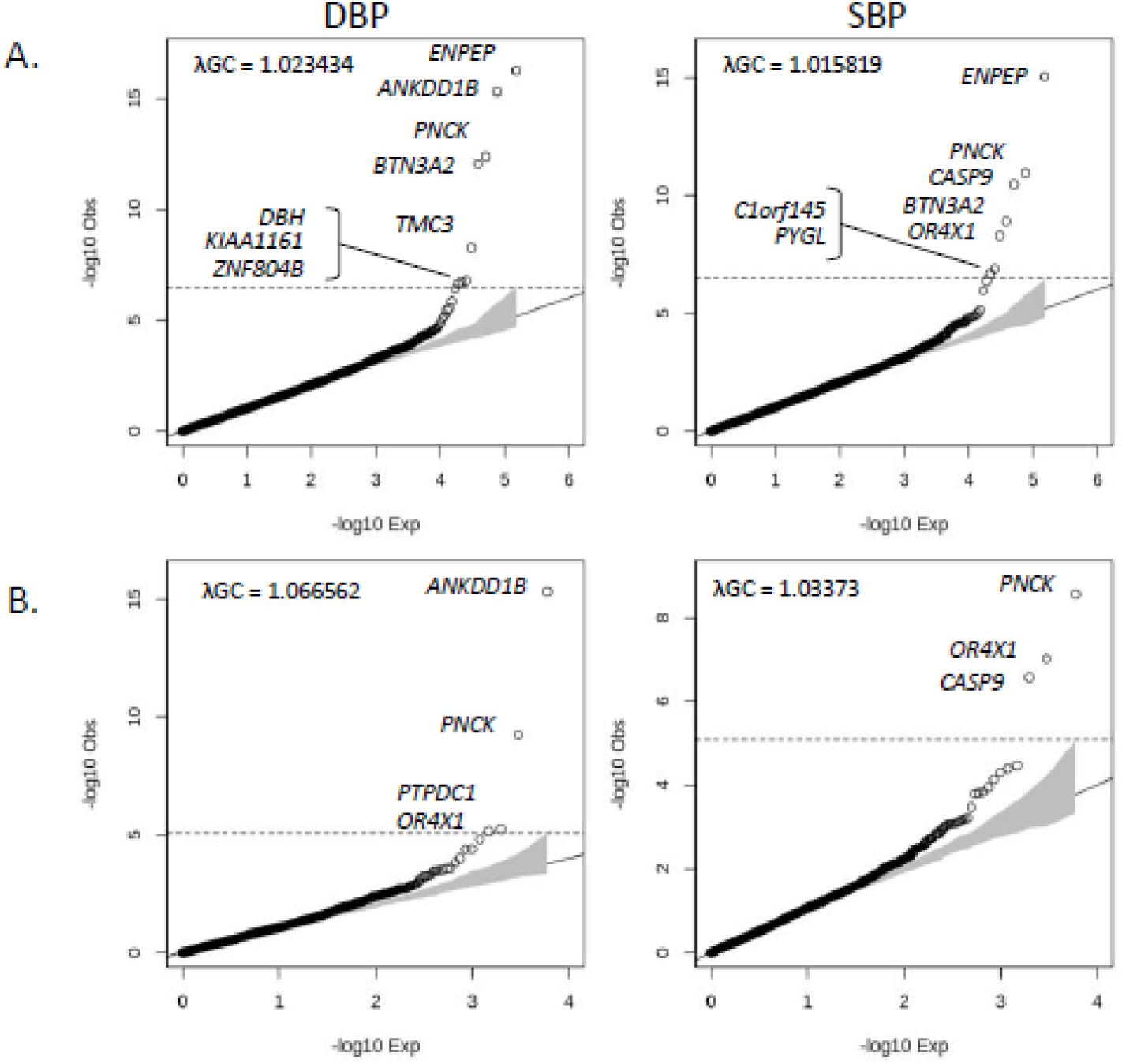
Quantile-quantile (Q-Q) plots for predicted loss-of function (pLoF) variant association results with diastolic blood pressure (DEP) and systolic blood pressure (SBPJ in the complete 500k set (N_sample_=452,385) using (A) an additive model (N_variant_= 152,840) and (**B**) a recessive model (N_variant_=5,955). In each plot, the dashed line represents the statistical significance threshold (additive model p-val≤3.27e-07; recessive model p-val≤8.40e-05) and the grey area is the 95% confidence interval.

**Supplementary Figure 4.**
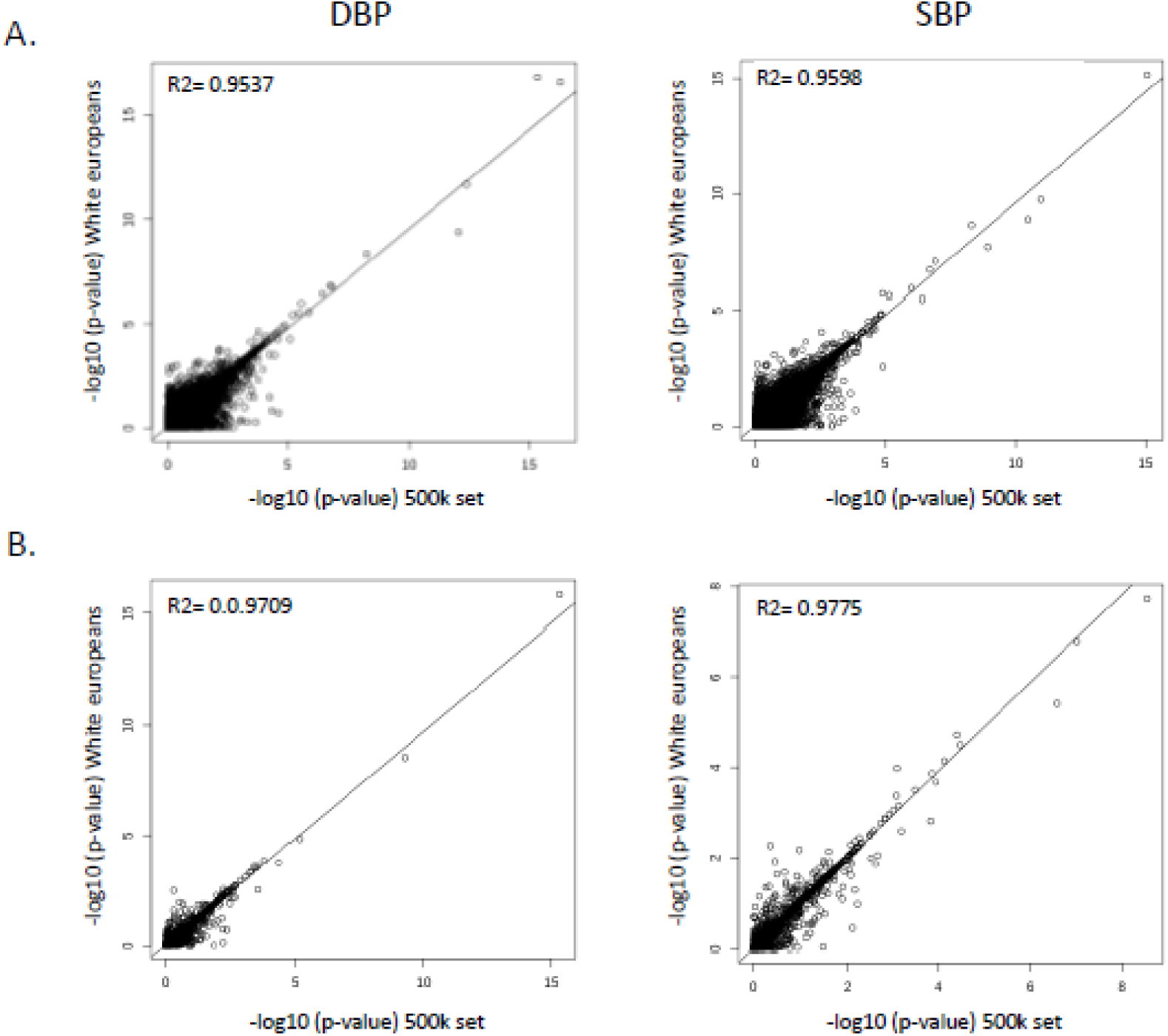
Comparison of single-variant association P-values obtained in the multi­ancestry 500k set (x-axis) and in the White British participants subset of the 500k set (N_sample_=428,381). (**A**) Additive and (**B**) recessive genetic models.

**Supplementary Figure 5.**
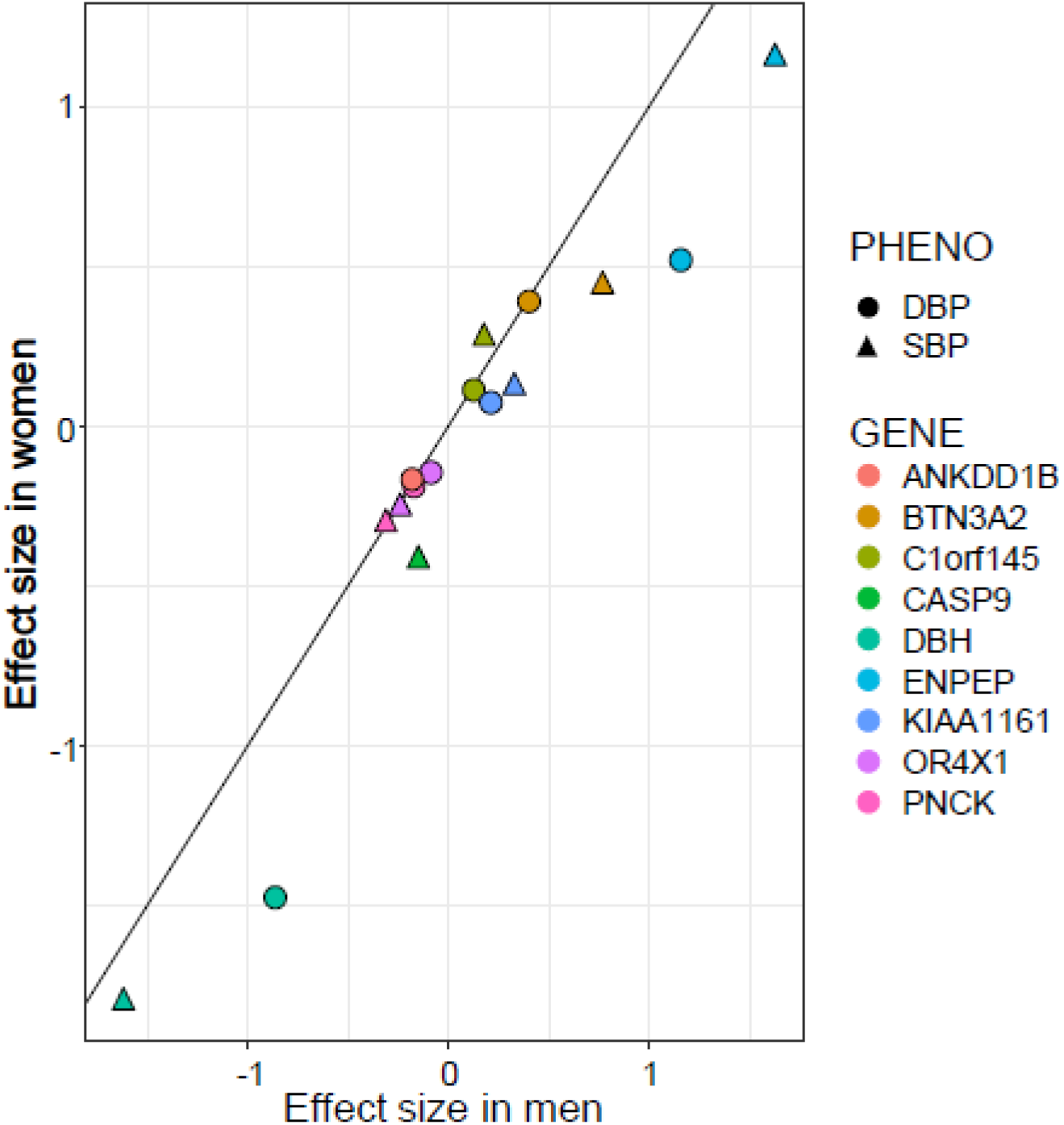
Comparison of effect sizes (in standardized mmHg) between men (x-axis) and women (x-axis) in the UK Biobank for each predicted loss-of-function (pLoF) variants associated with blood pressure. The differences are non-significant.

**Supplementary Figure 6.**
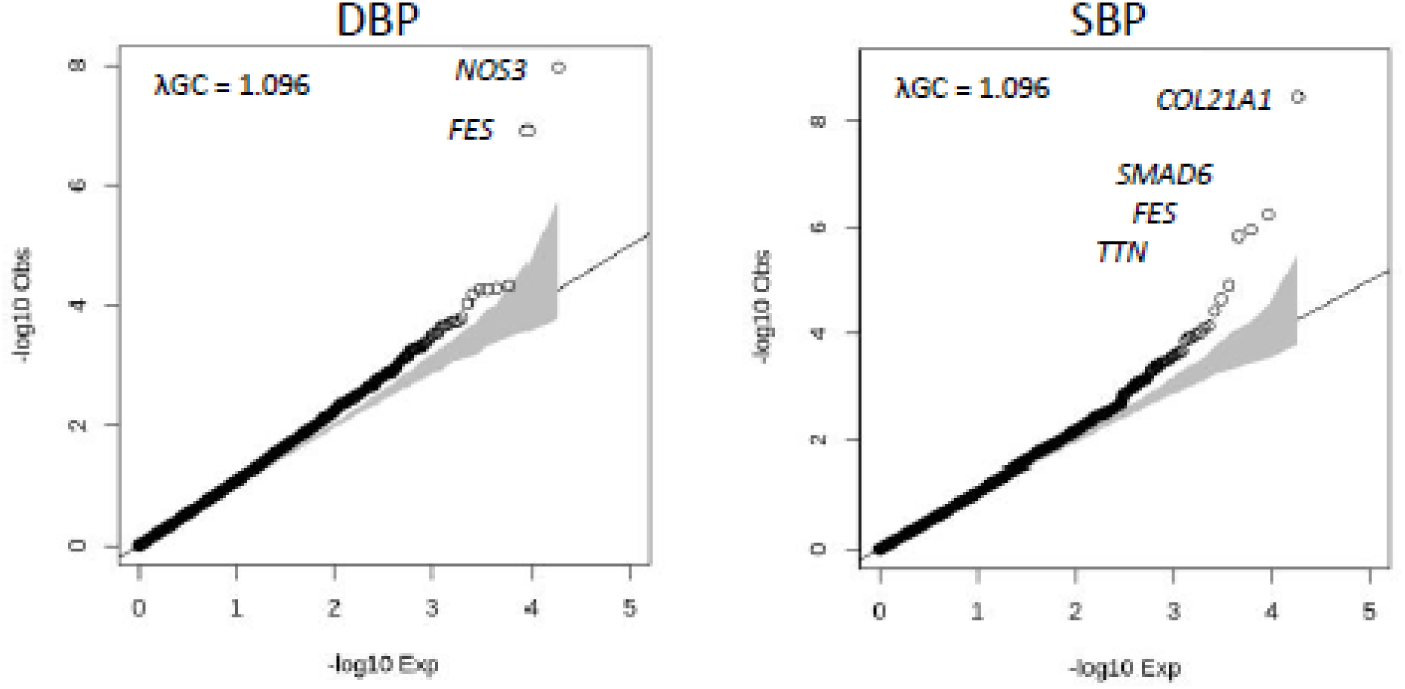
Quantile-quantile (Q-Q) plots for gene-based results with diastolic blood pressure (DBP) and systolic blood pressure (SBP) in the complete 500k set (N_sample_=452,385, N_gene_=18,436). We only tested predicted loss-of function (pLoF) variants with minor allele frequencies ≤0.1%. The grey area is the 95% confidence interval.

**Supplementary Figure 8.**
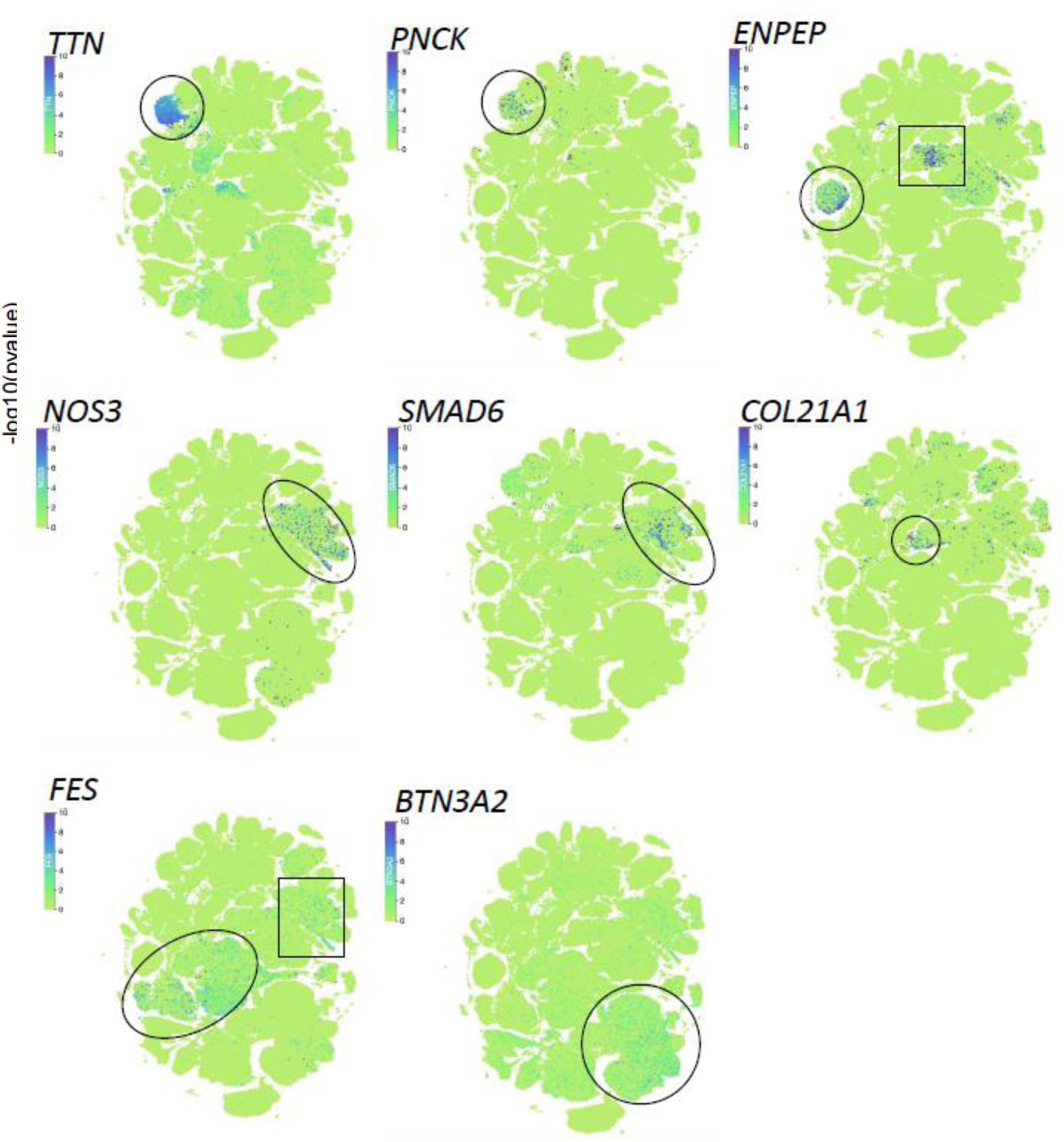
Uniform Manifold Approximation and Projection (UMAP) for 483,152 cells from 45 human tissues analyzed by the Tabula Sapiens Consortium (https://cellxgene.cziscience.com/). For each of the eight blood pressure (BP) genes with distinguishable expression profiles, we highlighted the main cell­type(s) based on annotations provided by Tabula Sapiens. *TTN* and *PNCK,* cardiomyocytes; *ENPEP,* kidney epithelial cells (circle) and smooth muscles/pericytes (rectangle); *NOS3* and *SMAE)6,* endothelial cells; *COL21A1,* smooth muscle cells, pericytes, and fibroblasts; FES, myeloid cells (oval) and endothelial cells (rectangle); BTN3A2, T-cells.

## References

1. Whelton PK, Carey RM, Aronow WS, et al. 2017 ACC/AHA/AAPA/ABC/ACPM/AGS/APhA/ASH/ASPC/NMA/PCNA Guideline for the Prevention, Detection, Evaluation, and Management of High Blood Pressure in Adults: Executive Summary: A Report of the American College of Cardiology/American Heart Association Task Force on Clinical Practice Guidelines. Hypertension. 2018;71(6):1269–1324. doi:10.1161/HYP.0000000000000066

2. Kearney PM, Whelton M, Reynolds K, Muntner P, Whelton PK, He J. Global burden of hypertension: analysis of worldwide data. Lancet. 2005;365(9455):217–223. doi:10.1016/S0140-6736(05)17741-1

3. NCD Risk Factor Collaboration (NCD-RisC). Worldwide trends in blood pressure from 1975 to 2015: a pooled analysis of 1479 population-based measurement studies with 19·1 million participants. Lancet. 2017;389(10064):37–55. doi:10.1016/S0140-6736(16)31919-5

4. Harrison DG, Coffman TM, Wilcox CS. Pathophysiology of Hypertension – the Mosaic Theory and Beyond. Circ Res. 2021;128(7):847–863. doi:10.1161/CIRCRESAHA.121.318082

5. Havlik RJ, Garrison RJ, Feinleib M, Kannel WB, Castelli WP, McNamara PM. Blood pressure aggregation in families. Am J Epidemiol. 1979;110(3):304–312. doi:10.1093/oxfordjournals.aje.a112815

6. Evangelou E, Warren HR, Mosen-Ansorena D, et al. Genetic analysis of over 1 million people identifies 535 new loci associated with blood pressure traits. Nature Genetics. 2018;50(10):1412–1425. doi:10.1038/s41588-018-0205-x

7. Giri A, Hellwege JN, Keaton JM, et al. Trans-ethnic association study of blood pressure determinants in over 750,000 individuals. Nature Genetics. 2019;51(1):51–62. doi:10.1038/s41588-018-0303-9

8. Surendran P, Feofanova EV, Lahrouchi N, et al. Discovery of rare variants associated with blood pressure regulation through meta-analysis of 1.3 million individuals. Nat Genet. 2020;52(12):1314–1332. doi:10.1038/s41588-020-00713-x

9. Sakaue S, Kanai M, Tanigawa Y, et al. A cross-population atlas of genetic associations for 220 human phenotypes. Nat Genet. 2021;53(10):1415–1424. doi:10.1038/s41588-021-00931-x

10. Mbatchou J, Barnard L, Backman J, et al. Computationally efficient whole-genome regression for quantitative and binary traits. Nat Genet. 2021;53(7):1097–1103. doi:10.1038/s41588-021-00870-7

11. Sollis E, Mosaku A, Abid A, et al. The NHGRI-EBI GWAS Catalog: knowledgebase and deposition resource. Nucleic Acids Res. 2023;51(D1):D977–D985. doi:10.1093/nar/gkac1010

12. Ochoa D, Hercules A, Carmona M, et al. The next-generation Open Targets Platform: reimagined, redesigned, rebuilt. Nucleic Acids Research. 2023;51(D1):D1353–D1359. doi:10.1093/nar/gkac1046

13. Huang D, Zhou Y, Yi X, et al. VannoPortal: multiscale functional annotation of human genetic variants for interrogating molecular mechanism of traits and diseases. Nucleic Acids Research. 2022;50(D1):D1408–D1416. doi:10.1093/nar/gkab853

14. Huang L, Rosen JD, Sun Q, et al. TOP-LD: A tool to explore linkage disequilibrium with TOPMed whole-genome sequence data. The American Journal of Human Genetics. 2022;109(6):1175–1181. doi:10.1016/j.ajhg.2022.04.006

15. Tabula Sapiens Consortium*, Jones RC, Karkanias J, et al. The Tabula Sapiens: A multiple-organ, single-cell transcriptomic atlas of humans. Science. 2022;376(6594):eabl4896. doi:10.1126/science.abl4896

16. Backman JD, Li AH, Marcketta A, et al. Exome sequencing and analysis of 454,787 UK Biobank participants. Nature. 2021;599(7886):628–634. doi:10.1038/s41586-021-04103-z

17. Arkes H. Once more unto the breach: the right to die--again. Issues Law Med. 1992;8(3):317–330.

18. Kim CH, Zabetian CP, Cubells JF, et al. Mutations in the dopamine beta-hydroxylase gene are associated with human norepinephrine deficiency. Am J Med Genet. 2002;108(2):140–147.

19. Gardner HP, Rajan JV, Ha SI, et al. Cloning, characterization, and chromosomal localization of Pnck, a Ca(2+)/calmodulin-dependent protein kinase. Genomics. 2000;63(2):279–288. doi:10.1006/geno.1999.6091

20. Brew O, Sullivan MHF, Woodman A. Comparison of Normal and Pre-Eclamptic Placental Gene Expression: A Systematic Review with Meta-Analysis. PLoS One. 2016;11(8):e0161504. doi:10.1371/journal.pone.0161504

21. Prasad AM, Morgan DA, Nuno DW, et al. Calcium/calmodulin-dependent kinase II inhibition in smooth muscle reduces angiotensin II-induced hypertension by controlling aortic remodeling and baroreceptor function. J Am Heart Assoc. 2015;4(6):e001949. doi:10.1161/JAHA.115.001949

22. Murthy S, Koval OM, Diaz JMR, et al. Endothelial CaMKII as a regulator of eNOS activity and NO-mediated vasoreactivity. PLOS ONE. 2017;12(10):e0186311. doi:10.1371/journal.pone.0186311

23. Fromont C, Atzori A, Kaur D, et al. Discovery of Highly Selective Inhibitors of Calmodulin-Dependent Kinases That Restore Insulin Sensitivity in the Diet-Induced Obesity in Vivo Mouse Model. J Med Chem. 2020;63(13):6784–6801. doi:10.1021/acs.jmedchem.9b01803

24. Hartiala JA, Han Y, Jia Q, et al. Genome-wide analysis identifies novel susceptibility loci for myocardial infarction. Eur Heart J. 2021;42(9):919–933. doi:10.1093/eurheartj/ehaa1040

25. Yakala GK, Cabrera-Fuentes HA, Crespo-Avilan GE, et al. FURIN Inhibition Reduces Vascular Remodeling and Atherosclerotic Lesion Progression in Mice. Arteriosclerosis, Thrombosis, and Vascular Biology. 2019;39(3):387–401. doi:10.1161/ATVBAHA.118.311903

26. Stolze LK, Conklin AC, Whalen MB, et al. Systems Genetics in Human Endothelial Cells Identifies Non-coding Variants Modifying Enhancers, Expression, and Complex Disease Traits. Am J Hum Genet. 2020;106(6):748–763. doi:10.1016/j.ajhg.2020.04.008

27. GTEx Consortium. The GTEx Consortium atlas of genetic regulatory effects across human tissues. Science. 2020;369(6509):1318–1330. doi:10.1126/science.aaz1776

28. Wünnemann F, Tadjo TF, Beaudoin M, Lalonde S, Lo KS, Lettre G. CRISPR perturbations at many coronary artery disease loci impair vascular endothelial cell functions. Published online February 11, 2021:2021.02.10.430527. doi:10.1101/2021.02.10.430527

29. Karczewski KJ, Solomonson M, Chao KR, et al. Systematic single-variant and gene-based association testing of thousands of phenotypes in 394,841 UK Biobank exomes. Cell Genom. 2022;2(9):100168. doi:10.1016/j.xgen.2022.100168

30. Minikel EV, Karczewski KJ, Martin HC, et al. Evaluating drug targets through human loss-of-function genetic variation. Nature. 2020;581(7809):459–464. doi:10.1038/s41586-020-2267-z

31. Lessard S, Manning AK, Low-Kam C, et al. Testing the role of predicted gene knockouts in human anthropometric trait variation. Human Molecular Genetics. 2016;25(10):2082–2092. doi:10.1093/hmg/ddw055

32. MacArthur DG, Balasubramanian S, Frankish A, et al. A systematic survey of loss-of-function variants in human protein-coding genes. Science. 2012;335(6070):823–828. doi:10.1126/science.1215040

33. Li AH, Morrison AC, Kovar C, et al. Analysis of loss-of-function variants and 20 risk factor phenotypes in 8,554 individuals identifies loci influencing chronic disease. Nat Genet. 2015;47(6):640–642. doi:10.1038/ng.3270

34. Lim ET, Würtz P, Havulinna AS, et al. Distribution and medical impact of loss-of-function variants in the Finnish founder population. PLoS Genet. 2014;10(7):e1004494. doi:10.1371/journal.pgen.1004494

35. Warren HR, Evangelou E, Cabrera CP, et al. Genome-wide association analysis identifies novel blood pressure loci and offers biological insights into cardiovascular risk. Nat Genet. 2017;49(3):403–415. doi:10.1038/ng.3768

36. Hoffmann TJ, Ehret GB, Nandakumar P, et al. Genome-wide association analyses using electronic health records identify new loci influencing blood pressure variation. Nat Genet. 2017;49(1):54–64. doi:10.1038/ng.3715

37. Kichaev G, Bhatia G, Loh PR, et al. Leveraging Polygenic Functional Enrichment to Improve GWAS Power. The American Journal of Human Genetics. 2019;104(1):65–75. doi:10.1016/j.ajhg.2018.11.008

38. Kato N, Takeuchi F, Tabara Y, et al. Meta-analysis of genome-wide association studies identifies common variants associated with blood pressure variation in east Asians. Nat Genet. 2011;43(6):531–538. doi:10.1038/ng.834

39. Barton AR, Sherman MA, Mukamel RE, Loh PR. Whole-exome imputation within UK Biobank powers rare coding variant association and fine-mapping analyses. Nat Genet. 2021;53(8):1260–1269. doi:10.1038/s41588-021-00892-1

40. Klimentidis YC, Arora A, Newell M, et al. Phenotypic and Genetic Characterization of Lower LDL Cholesterol and Increased Type 2 Diabetes Risk in the UK Biobank. Diabetes. 2020;69(10):2194–2205. doi:10.2337/db19-1134

41. Willer CJ, Schmidt EM, Sengupta S, et al. Discovery and refinement of loci associated with lipid levels. Nat Genet. 2013;45(11):1274–1283. doi:10.1038/ng.2797

42. Teslovich TM, Musunuru K, Smith AV, et al. Biological, clinical and population relevance of 95 loci for blood lipids. Nature. 2010;466(7307):707–713. doi:10.1038/nature09270

43. Locke AE, Kahali B, Berndt SI, et al. Genetic studies of body mass index yield new insights for obesity biology. Nature. 2015;518(7538):197–206. doi:10.1038/nature14177

44. UK Biobank. Neale lab. Accessed November 28, 2022. http://www.nealelab.is/uk-biobank

45. Chen MH, Raffield LM, Mousas A, et al. Trans-ethnic and Ancestry-Specific Blood-Cell Genetics in 746,667 Individuals from 5 Global Populations. Cell. 2020;182(5):1198–1213.e14. doi:10.1016/j.cell.2020.06.045

46. Vuckovic D, Bao EL, Akbari P, et al. The Polygenic and Monogenic Basis of Blood Traits and Diseases. Cell. 2020;182(5):1214–1231.e11. doi:10.1016/j.cell.2020.08.008

47. Ripatti P, Rämö JT, Mars NJ, et al. Polygenic Hyperlipidemias and Coronary Artery Disease Risk. Circulation: Genomic and Precision Medicine. 2020;13(2):e002725. doi:10.1161/CIRCGEN.119.002725

48. Richardson TG, Sanderson E, Palmer TM, et al. Evaluating the relationship between circulating lipoprotein lipids and apolipoproteins with risk of coronary heart disease: A multivariable Mendelian randomisation analysis. PLoS Med. 2020;17(3):e1003062. doi:10.1371/journal.pmed.1003062

49. Sinnott-Armstrong N, Tanigawa Y, Amar D, et al. Genetics of 35 blood and urine biomarkers in the UK Biobank. Nat Genet. 2021;53(2):185–194. doi:10.1038/s41588-020-00757-z

50. Pulit SL, Stoneman C, Morris AP, et al. Meta-analysis of genome-wide association studies for body fat distribution in 694 649 individuals of European ancestry. Human Molecular Genetics. 2019;28(1):166–174. doi:10.1093/hmg/ddy327

51. Hoffmann TJ, Theusch E, Haldar T, et al. A large electronic-health-record-based genome-wide study of serum lipids. Nat Genet. 2018;50(3):401–413. doi:10.1038/s41588-018-0064-5

52. Klarin D, Damrauer SM, Cho K, et al. Genetics of blood lipids among ∼300,000 multi-ethnic participants of the Million Veteran Program. Nat Genet. 2018;50(11):1514–1523. doi:10.1038/s41588-018-0222-9

53. Kurki MI, Karjalainen J, Palta P, et al. FinnGen: Unique genetic insights from combining isolated population and national health register data. Published online March 6, 2022:2022.03.03.22271360. doi:10.1101/2022.03.03.22271360

54. Hu Y, Stilp AM, McHugh CP, et al. Whole-genome sequencing association analysis of quantitative red blood cell phenotypes: The NHLBI TOPMed program. The American Journal of Human Genetics. 2021;108(5):874–893. doi:10.1016/j.ajhg.2021.04.003

55. Dönertaş HM, Fabian DK, Fuentealba M, Partridge L, Thornton JM. Common genetic associations between age-related diseases. Nat Aging. 2021;1(4):400–412. doi:10.1038/s43587-021-00051-5

56. Feofanova EV, Chen H, Dai Y, et al. A Genome-wide Association Study Discovers 46 Loci of the Human Metabolome in the Hispanic Community Health Study/Study of Latinos. The American Journal of Human Genetics. 2020;107(5):849–863. doi:10.1016/j.ajhg.2020.09.003

57. Schlosser P, Li Y, Sekula P, et al. Genetic studies of urinary metabolites illuminate mechanisms of detoxification and excretion in humans. Nat Genet. 2020;52(2):167–176. doi:10.1038/s41588-019-0567-8

58. Morris JA, Kemp JP, Youlten SE, et al. An atlas of genetic influences on osteoporosis in humans and mice. Nat Genet. 2019;51(2):258–266. doi:10.1038/s41588-018-0302-x

59. Kim SK. Identification of 613 new loci associated with heel bone mineral density and a polygenic risk score for bone mineral density, osteoporosis and fracture. PLOS ONE. 2018;13(7):e0200785. doi:10.1371/journal.pone.0200785

60. Kemp JP, Morris JA, Medina-Gomez C, et al. Identification of 153 new loci associated with heel bone mineral density and functional involvement of GPC6 in osteoporosis. Nat Genet. 2017;49(10):1468–1475. doi:10.1038/ng.3949

61. Gill D, Cameron AC, Burgess S, et al. Urate, Blood Pressure, and Cardiovascular Disease. Hypertension. 2021;77(2):383–392. doi:10.1161/HYPERTENSIONAHA.120.16547

62. Tin A, Marten J, Halperin Kuhns VL, et al. Target genes, variants, tissues and transcriptional pathways influencing human serum urate levels. Nat Genet. 2019;51(10):1459–1474. doi:10.1038/s41588-019-0504-x

63. Kanai M, Akiyama M, Takahashi A, et al. Genetic analysis of quantitative traits in the Japanese population links cell types to complex human diseases. Nat Genet. 2018;50(3):390–400. doi:10.1038/s41588-018-0047-6

64. Pirruccello JP, Bick A, Wang M, et al. Analysis of cardiac magnetic resonance imaging in 36,000 individuals yields genetic insights into dilated cardiomyopathy. Nat Commun. 2020;11(1):2254. doi:10.1038/s41467-020-15823-7

65. Tadros R, Francis C, Xu X, et al. Shared genetic pathways contribute to risk of hypertrophic and dilated cardiomyopathies with opposite directions of effect. Nat Genet. 2021;53(2):128–134. doi:10.1038/s41588-020-00762-2

66. Harper AR, Goel A, Grace C, et al. Common genetic variants and modifiable risk factors underpin hypertrophic cardiomyopathy susceptibility and expressivity. Nat Genet. 2021;53(2):135–142. doi:10.1038/s41588-020-00764-0

67. Kachuri L, Jeon S, DeWan AT, et al. Genetic determinants of blood-cell traits influence susceptibility to childhood acute lymphoblastic leukemia. The American Journal of Human Genetics. 2021;108(10):1823–1835. doi:10.1016/j.ajhg.2021.08.004

68. Astle WJ, Elding H, Jiang T, et al. The Allelic Landscape of Human Blood Cell Trait Variation and Links to Common Complex Disease. Cell. 2016;167(5):1415–1429.e19. doi:10.1016/j.cell.2016.10.042

69. Wood AR, Esko T, Yang J, et al. Defining the role of common variation in the genomic and biological architecture of adult human height. Nat Genet. 2014;46(11):1173–1186. doi:10.1038/ng.3097

70. Lango Allen H, Estrada K, Lettre G, et al. Hundreds of variants clustered in genomic loci and biological pathways affect human height. Nature. 2010;467(7317):832–838. doi:10.1038/nature09410

71. Berndt SI, Gustafsson S, Mägi R, et al. Genome-wide meta-analysis identifies 11 new loci for anthropometric traits and provides insights into genetic architecture. Nat Genet. 2013;45(5):501–512. doi:10.1038/ng.2606

72. Ehret GB, Ferreira T, Chasman DI, et al. The genetics of blood pressure regulation and its target organs from association studies in 342,415 individuals. Nat Genet. 2016;48(10):1171–1184. doi:10.1038/ng.3667

73. Sabater-Lleal M, Huffman JE, de Vries PS, et al. Genome-Wide Association Transethnic Meta-Analyses Identifies Novel Associations Regulating Coagulation Factor VIII and von Willebrand Factor Plasma Levels. Circulation. 2019;139(5):620–635. doi:10.1161/CIRCULATIONAHA.118.034532

74. Gilly A, Park YC, Png G, et al. Whole-genome sequencing analysis of the cardiometabolic proteome. Nat Commun. 2020;11(1):6336. doi:10.1038/s41467-020-20079-2

75. Chen VL, Du X, Chen Y, et al. Genome-wide association study of serum liver enzymes implicates diverse metabolic and liver pathology. Nat Commun. 2021;12(1):816. doi:10.1038/s41467-020-20870-1

76. Li J, Gui L, Wu C, et al. Genome-wide association study on serum alkaline phosphatase levels in a Chinese population. BMC Genomics. 2013;14(1):684. doi:10.1186/1471-2164-14-684

77. Prins BP, Kuchenbaecker KB, Bao Y, et al. Genome-wide analysis of health-related biomarkers in the UK Household Longitudinal Study reveals novel associations. Sci Rep. 2017;7(1):11008. doi:10.1038/s41598-017-10812-1

78. Sliz E, Kalaoja M, Ahola-Olli A, et al. Genome-wide association study identifies seven novel loci associating with circulating cytokines and cell adhesion molecules in Finns. J Med Genet. 2019;56(9):607–616. doi:10.1136/jmedgenet-2018-105965

79. Teng MS, Hsu LA, Wu S, Tzeng IS, Chou HH, Ko YL. Genome-wide association study revealed novel candidate gene loci associated with soluble E-selectin levels in a Taiwanese population. Atherosclerosis. 2021;337:18–26. doi:10.1016/j.atherosclerosis.2021.10.006

80. Paterson AD, Lopes-Virella MF, Waggott D, et al. Genome-Wide Association Identifies the ABO Blood Group as a Major Locus Associated With Serum Levels of Soluble E-Selectin. Arteriosclerosis, Thrombosis, and Vascular Biology. 2009;29(11):1958–1967. doi:10.1161/ATVBAHA.109.192971

81. Folkersen L, Gustafsson S, Wang Q, et al. Genomic and drug target evaluation of 90 cardiovascular proteins in 30,931 individuals. Nat Metab. 2020;2(10):1135–1148. doi:10.1038/s42255-020-00287-2

82. Roselli C, Chaffin MD, Weng LC, et al. Multi-ethnic genome-wide association study for atrial fibrillation. Nat Genet. 2018;50(9):1225–1233. doi:10.1038/s41588-018-0133-9

83. Nielsen JB, Thorolfsdottir RB, Fritsche LG, et al. Biobank-driven genomic discovery yields new insight into atrial fibrillation biology. Nat Genet. 2018;50(9):1234–1239. doi:10.1038/s41588-018-0171-3

84. Christophersen IE, Rienstra M, Roselli C, et al. Large-scale analyses of common and rare variants identify 12 new loci associated with atrial fibrillation. Nat Genet. 2017;49(6):946–952. doi:10.1038/ng.3843

85. Ellinor PT, Lunetta KL, Albert CM, et al. Meta-analysis identifies six new susceptibility loci for atrial fibrillation. Nat Genet. 2012;44(6):670–675. doi:10.1038/ng.2261

86. Benjamin EJ, Rice KM, Arking DE, et al. Variants in ZFHX3 are associated with atrial fibrillation in individuals of European ancestry. Nat Genet. 2009;41(8):879–881. doi:10.1038/ng.416

87. Ishigaki K, Akiyama M, Kanai M, et al. Large-scale genome-wide association study in a Japanese population identifies novel susceptibility loci across different diseases. Nat Genet. 2020;52(7):669–679. doi:10.1038/s41588-020-0640-3

88. Malik R, Chauhan G, Traylor M, et al. Multiancestry genome-wide association study of 520,000 subjects identifies 32 loci associated with stroke and stroke subtypes. Nat Genet. 2018;50(4):524–537. doi:10.1038/s41588-018-0058-3

89. Dichgans M, Malik R, König IR, et al. Shared Genetic Susceptibility to Ischemic Stroke and Coronary Artery Disease. Stroke. 2014;45(1):24–36. doi:10.1161/STROKEAHA.113.002707

90. Ehret GB, Munroe PB, Rice KM, et al. Genetic variants in novel pathways influence blood pressure and cardiovascular disease risk. Nature. 2011;478(7367):103–109. doi:10.1038/NATURE10405

91. Gudjonsson A, Gudmundsdottir V, Axelsson GT, et al. A genome-wide association study of serum proteins reveals shared loci with common diseases. Nat Commun. 2022;13(1):480. doi:10.1038/s41467-021-27850-z

92. Emilsson V, Ilkov M, Lamb JR, et al. Co-regulatory networks of human serum proteins link genetics to disease. Science. 2018;361(6404):769–773. doi:10.1126/science.aaq1327

93. Sun W, Kechris K, Jacobson S, et al. Common Genetic Polymorphisms Influence Blood Biomarker Measurements in COPD. PLOS Genetics. 2016;12(8):e1006011. doi:10.1371/journal.pgen.1006011

94. Ruffieux H, Carayol J, Popescu R, et al. A fully joint Bayesian quantitative trait locus mapping of human protein abundance in plasma. PLOS Computational Biology. 2020;16(6):e1007882. doi:10.1371/journal.pcbi.1007882

95. Carayol J, Chabert C, Di Cara A, et al. Protein quantitative trait locus study in obesity during weight-loss identifies a leptin regulator. Nat Commun. 2017;8(1):2084. doi:10.1038/s41467-017-02182-z

96. Ahola-Olli AV, Würtz P, Havulinna AS, et al. Genome-wide Association Study Identifies 27 Loci Influencing Concentrations of Circulating Cytokines and Growth Factors. The American Journal of Human Genetics. 2017;100(1):40–50. doi:10.1016/j.ajhg.2016.11.007

97. Akiyama M, Ishigaki K, Sakaue S, et al. Characterizing rare and low-frequency height-associated variants in the Japanese population. Nat Commun. 2019;10(1):4393. doi:10.1038/s41467-019-12276-5

98. Liu C, Kraja AT, Smith JA, et al. Meta-analysis identifies common and rare variants influencing blood pressure and overlapping with metabolic trait loci. Nature genetics. 2016;48(10):1162–1170. doi:10.1038/NG.3660

99. Pei YF, Liu YZ, Yang XL, et al. The genetic architecture of appendicular lean mass characterized by association analysis in the UK Biobank study. Commun Biol. 2020;3(1):1–13. doi:10.1038/s42003-020-01334-0

100. Hernandez Cordero AI, Gonzales NM, Parker CC, et al. Genome-wide Associations Reveal Human-Mouse Genetic Convergence and Modifiers of Myogenesis, CPNE1 and STC2. Am J Hum Genet. 2019;105(6):1222–1236. doi:10.1016/j.ajhg.2019.10.014

101. Bihlmeyer NA, Brody JA, Smith AV, et al. ExomeChip-Wide Analysis of 95 626 Individuals Identifies 10 Novel Loci Associated With QT and JT Intervals. Circulation: Genomic and Precision Medicine. 2018;11(1):e001758. doi:10.1161/CIRCGEN.117.001758

102. Arking DE, Pulit SL, Crotti L, et al. Genetic association study of QT interval highlights role for calcium signaling pathways in myocardial repolarization. Nat Genet. 2014;46(8):826–836. doi:10.1038/ng.3014

103. van Duijvenboden S, Ramírez J, Young WJ, et al. Genomic and pleiotropic analyses of resting QT interval identifies novel loci and overlap with atrial electrical disorders. Human Molecular Genetics. 2021;30(24):2513–2523. doi:10.1093/hmg/ddab197

104. van Setten J, Verweij N, Mbarek H, et al. Genome-wide association meta-analysis of 30,000 samples identifies seven novel loci for quantitative ECG traits. Eur J Hum Genet. 2019;27(6):952–962. doi:10.1038/s41431-018-0295-z

105. Newton-Cheh C, Eijgelsheim M, Rice KM, et al. Common variants at ten loci influence QT interval duration in the QTGEN Study. Nat Genet. 2009;41(4):399–406. doi:10.1038/ng.364

106. Pfeufer A, Sanna S, Arking DE, et al. Common variants at ten loci modulate the QT interval duration in the QTSCD Study. Nat Genet. 2009;41(4):407–414. doi:10.1038/ng.362

107. Méndez-Giráldez R, Gogarten SM, Below JE, et al. GWAS of the electrocardiographic QT interval in Hispanics/Latinos generalizes previously identified loci and identifies population-specific signals. Sci Rep. 2017;7(1):17075. doi:10.1038/s41598-017-17136-0

108. Wojcik GL, Graff M, Nishimura KK, et al. Genetic analyses of diverse populations improves discovery for complex traits. Nature. 2019;570(7762):514–518. doi:10.1038/s41586-019-1310-4

109. German CA, Sinsheimer JS, Klimentidis YC, Zhou H, Zhou JJ. Ordered multinomial regression for genetic association analysis of ordinal phenotypes at Biobank scale. Genetic Epidemiology. 2020;44(3):248–260. doi:10.1002/gepi.22276

110. Pilling LC, Atkins JL, Duff MO, et al. Red blood cell distribution width: Genetic evidence for aging pathways in 116,666 volunteers. PLOS ONE. 2017;12(9):e0185083. doi:10.1371/journal.pone.0185083

111. Kato N, Loh M, Takeuchi F, et al. Trans-ancestry genome-wide association study identifies 12 genetic loci influencing blood pressure and implicates a role for DNA methylation. Nature Genetics. 2015;47(11):1282–1293. doi:10.1038/ng.3405

112. Pietzner M, Wheeler E, Carrasco-Zanini J, et al. Genetic architecture of host proteins involved in SARS-CoV-2 infection. Nat Commun. 2020;11(1):6397. doi:10.1038/s41467-020-19996-z

113. Eppinga RN, Hagemeijer Y, Burgess S, et al. Identification of genomic loci associated with resting heart rate and shared genetic predictors with all-cause mortality. Nat Genet. 2016;48(12):1557–1563. doi:10.1038/ng.3708

114. Ramírez J, Duijvenboden S van, Ntalla I, et al. Thirty loci identified for heart rate response to exercise and recovery implicate autonomic nervous system. Nat Commun. 2018;9(1):1947. doi:10.1038/s41467-018-04148-1

115. den Hoed M, Eijgelsheim M, Esko T, et al. Identification of heart rate–associated loci and their effects on cardiac conduction and rhythm disorders. Nat Genet. 2013;45(6):621–631. doi:10.1038/ng.2610

116. Demenais F, Margaritte-Jeannin P, Barnes KC, et al. Multiancestry association study identifies new asthma risk loci that colocalize with immune-cell enhancer marks. Nat Genet. 2018;50(1):42–53. doi:10.1038/s41588-017-0014-7

117. Han Y, Jia Q, Jahani PS, et al. Genome-wide analysis highlights contribution of immune system pathways to the genetic architecture of asthma. Nat Commun. 2020;11(1):1776. doi:10.1038/s41467-020-15649-3

118. Johansson Å, Rask-Andersen M, Karlsson T, Ek WE. Genome-wide association analysis of 350 000 Caucasians from the UK Biobank identifies novel loci for asthma, hay fever and eczema. Human Molecular Genetics. 2019;28(23):4022–4041. doi:10.1093/hmg/ddz175

119. Moffatt MF, Kabesch M, Liang L, et al. Genetic variants regulating ORMDL3 expression contribute to the risk of childhood asthma. Nature. 2007;448(7152):470–473. doi:10.1038/nature06014

120. Valette K, Li Z, Bon-Baret V, et al. Prioritization of candidate causal genes for asthma in susceptibility loci derived from UK Biobank. Commun Biol. 2021;4(1):1–15. doi:10.1038/s42003-021-02227-6

121. Olafsdottir TA, Theodors F, Bjarnadottir K, et al. Eighty-eight variants highlight the role of T cell regulation and airway remodeling in asthma pathogenesis. Nat Commun. 2020;11(1):393. doi:10.1038/s41467-019-14144-8

122. Ferreira MAR, Mathur R, Vonk JM, et al. Genetic Architectures of Childhood- and Adult-Onset Asthma Are Partly Distinct. The American Journal of Human Genetics. 2019;104(4):665–684. doi:10.1016/j.ajhg.2019.02.022

123. Zhu Z, Lee PH, Chaffin MD, et al. A genome-wide cross-trait analysis from UK Biobank highlights the shared genetic architecture of asthma and allergic diseases. Nat Genet. 2018;50(6):857–864. doi:10.1038/s41588-018-0121-0

124. Pickrell JK, Berisa T, Liu JZ, Ségurel L, Tung JY, Hinds DA. Detection and interpretation of shared genetic influences on 42 human traits. Nat Genet. 2016;48(7):709–717. doi:10.1038/ng.3570

