## Supplemental results for "Association analyses of predicted loss-of-function variants prioritized 15 genes as blood pressure regulators"

### SUPPLEMENTARY TEXT

#### Known blood pressure (BP) regulators: *ENPEP*, *NOS3* and *DBH*

Several genes that had a LoF variant significantly associated to SBP or DBP were already known to be involved in BP regulation. Indeed, we highlighted *ENPEP*, which encodes glutamyl aminopeptidase, an enzyme implicated in angiotensin II degradation, and *NOS3*, the gene for eNOS, the enzyme that produces nitric oxide in endothelial cells, a vasodilator. A *DBH* LoF variant also seems to decrease BP, consistent with the observation of hypotension induced by *DBH* deficiency in human and mouse[1–3]. *DBH* encodes the dopamine beta-hydroxylase which catalyzed dopamine to norepinephrine conversion, a transmitter that maintains the heart rate. This gene is a promising drug target, and is currently under investigation for the development of new antihypertensive drug[4,5].

#### Gene with a role in renal functions: *CASP9*

*CASP9* frameshift variant rs2234723 is associated with SBP variation in our analysis and was only recently associated with cystatin C and creatinine levels[6]. This gene encodes caspase 9, a trigger of intrinsic apoptosis that may also be implicated in autophagy regulation. *CASP9* is associated with several serum metabolite levels, glomerular filtration rate and bone mineral density in other studies[6–9], and seems to play a causal role in kidney disease development[10].

#### Genes with roles in the cardiovascular systems: *SMAD6*, *COL21A1* and *TTN*

*SMAD6* is known to be mainly implicated in cardiovascular diseases, craniosynostosis and radioulnar synostosis[11–14]. It encodes SMAD family member 6 and is involved in the bone morphogenetic protein signalling pathway. In the heart, this gene is expressed in endocardial cells, adipocytes and endothelial cells of the left ventricle, and is downregulated in the latest in dilated and hypertrophic cardiomyopathy[15]. Recently, it has been shown that *SMAD6* plays a key role in endothelial cell junctions, vascular development, and homeostasis[16,17], which may explain its effect on BP.

A rare variant in *COL21A1* had already been associated with pulse pressure[18–20]. It encodes alpha-1 chain precursor of type XX1 collagen, and is expressed in several tissues, including the heart and aorta. In a pathological context, this gene is over-expressed in cardiomyocytes, fibroblasts and vascular smooth muscle cells of the left ventricle of dilated and hypertrophic cardiomyopathy[15]. It is also up-regulated in tibial artery, skeletal muscle and subcutaneous adipose tissues of hypertensive patients[21]. *TTN* encodes the giant muscle protein titin. Mutations in this gene have been linked to dilated cardiomyopathy[22], in particular truncating mutations located in the A-band of titin (as opposed to the Z-disk and M-band regions)[23].

#### Genes with unclear functions in BP regulation: *BTN3A2, ANKDD1B, OR4X1, TMC3, C1orf145 [OBSCN-AS1], KIAA1161 [MYORG]*

<u>*BTN3A2.*</u> The *BTN3A2* splice donor variant rs58367598 is associated with both SBP and DBP variation in our study and in another genetic study[24], as well as with red blood cell traits[6,25,26]. This gene encodes butyrophilin subfamily 3 member A2 and is implicated in the adaptive immune

system. This pLoF variant is also associated with indoxyl sulfate levels in the blood[27], and this metabolite is known to affect arterial blood pressure via peripheral and central mechanisms dependent on serotonin signaling in the rat[28,29]. The same variant is also linked with glutamine[27], an inhibitor of endothelial NO synthesis[30].

*ANKDD1B.* The common stop-gained variant rs34358 in *ANKDD1B* was highlighted by our analysis. This variant has already been associated with a wide range of traits, including type 2 diabetes, migraine[31], dyslipidemia, hypercholesterolemia, and blood and immune cells traits[6,32–34]. *ANKDD1B* encodes ankyrin repeat and death domain containing 1B, a protein that belongs to a family known to mediate protein-protein interactions[35] and implicated in various cellular processes. This gene is suspected of being implicated in ankylosing spondylitis[36], and a cross-trait GWAS meta-analysis has shown a plausible shared role of the gene between migraine and major depressive disorders[37], and migraine and blood pressure[38].The gene was recently associated with DBP[6,20], but its role in BP regulation has never been studied. However, this protein truncating variant seems to mainly affect the death domain of the protein, and lowers BP. This may be consistent with the over-expression of the gene in adipose tissue of hypertensive patients[21]. The pLoF variant may therefore be protective of high BP.

*OR4X1.* A common stop gained variant in *OR4X1* seems to lower SBP measurement. rs10838851 is associated with sex hormone-binding globulin levels, heel bone mineral density, DBP, and total testosterone levels. It is also linked to recurrent pregnancy lost in Korean women[39] and to the ratio of the forced expiratory volume in the first second and the forced vital capacity values of pulmonary function test[40]. At the gene level, it is linked with HDL cholesterol level, lymphocyte count, cardiomyopathy and blood pressure phenotypes. Besides GWAS results, little is known about this gene, and its expression across tissues remains elusive as it is not detected at RNA or protein level. However, it belongs to the family of olfactory receptor, and it has been demonstrated during this last decade that they are implicated in a wide range of biological processes, including BP regulation[41].

*TMC3.* A *TMC3* stop gained variant (rs150843673, MAF=22%) is associated with increased DBP. This gene encodes transmembrane channel like 3 and is also associated with height, pro-interleukin 16 levels, lymphocytes count and red blood cell traits.

*C1orf145(OBSCN-AS1).* The pLoF variant rs11800309 is usually annotated as a *OBSCN* variant, a gene located near the variant but without overlapping it. However, it is predicted to introduce a stop codon in the last exon of the antisense gene, *C1orf145* (*OBSCN-AS1*). Nothing is known about the protein encoded by this antisense RNA.

*KIAA1161(MYORG).* The common stop gained variant located in *KIAA1161*(*MYORG*) has never been associated with BP variation, but with immature reticulocyte fraction. This putative gene encodes myogenesis regulating glycosidase. It has been associated with serum levels of protein IL11RA and various blood cell traits.

**Supplementary Table 1.** Demographic information of the UK Biobank for this blood pressure genetic project. We used ancestry as defined in the UK Biobank dataset. Diastolic blood pressure (DBP) and systolic blood pressure (SBP) are expressed in millimeters of mercury (mmHg).

| | 200k WES set | 300k WES set | 450k WES set | Whole 500k cohort |
|---|---|---|---|---|
| White (sample size) | 188,181 | 240,284 | 428,381 | 472,542 |
| Black (sample size) | 3,222 | 3,884 | 7,104 | 8050 |
| Asian (sample size) | 4,897 | 5,375 | 10,271 | 11,437 |
| Mixed (sample size) | 1,305 | 1,339 | 2,642 | 2,961 |
| Other (sample size) | 2,997 | 3,314 | 6,311 | 7,421 |
| Anti-hypertension drugs (sample size) | 41,409 | 54,571 | 95,963 | 105,483 |
| DBP after correction for blood pressure lowering drugs, in mmHg (mean;median;sd) | 84.23; 83.5;11.31 | 84.43;84;11.3 | 84.34;83.5;11.30 | 84.33;83.5;20.8 |
| SBP after correction for blood pressure lowering drugs, in mmHg (mean;median;sd) | 140.72;138.5;20.64 | 141.21;139;20.73 | 141;139;20.70 | 141;139;20.75 |
| Male (sample size) | 90,141 | 118,009 | 208,120 | 229,084 |
| Female (sample size) | 110,461 | 136,187 | 246,589 | 273,327 |
| Age, years (mean;median;sd) | 56.46;58;8.1 | 56.59;58;8.09 | 56.5;58;8.09 | 56.5;58;8.09 |
| Total (sample size) | 200,602 | 254,196 | 454,709 | 502,412 |

References


1. Robertson D, Haile V, Perry SE, Robertson RM, Phillips JA, Biaggioni I. Dopamine beta-hydroxylase deficiency. A genetic disorder of cardiovascular regulation. *Hypertension*. 1991;18:1–8.

2. Swoap SJ, Weinshenker D, Palmiter RD, Garber G. Dbh(-/-) mice are hypotensive, have altered circadian rhythms, and have abnormal responses to dieting and stress. *Am J Physiol Regul Integr Comp Physiol*. 2004;286:R108-113.

3. Pravenec M, Landa V, Zídek V, Mlejnek P, Šilhavý J, Mir SA, Vaingankar SM, Wang J, Kurtz TW. Effects of transgenic expression of dopamine beta hydroxylase (Dbh) gene on blood pressure in spontaneously hypertensive rats. *Physiol Res*. 2016;65:1039–1044.

4. Catelas DN, Serrão MP, Soares-Da-Silva P. Effects of nepicastat upon dopamine-β-hydroxylase activity and dopamine and norepinephrine levels in the rat left ventricle, kidney, and adrenal gland. *Clinical and Experimental Hypertension*. 2020;42:118–125.

5. Dey SK, Saini M, Prabhakar P, Kundu S. Dopamine β hydroxylase as a potential drug target to combat hypertension. *Expert Opin Investig Drugs*. 2020;29:1043–1057.

6. Barton AR, Sherman MA, Mukamel RE, Loh P-R. Whole-exome imputation within UK Biobank powers rare coding variant association and fine-mapping analyses. *Nat Genet*. 2021;53:1260–1269.

7. Schlosser P, Li Y, Sekula P, Raffler J, Grundner-Culemann F, Pietzner M, Cheng Y, Wuttke M, Steinbrenner I, Schultheiss UT, et al. Genetic studies of urinary metabolites illuminate mechanisms of detoxification and excretion in humans. *Nat Genet*. 2020;52:167–176.

8. Morris JA, Kemp JP, Youlten SE, Laurent L, Logan JG, Chai RC, Vulpescu NA, Forgetta V, Kleinman A, Mohanty ST, et al. An atlas of genetic influences on osteoporosis in humans and mice. *Nat Genet*. 2019;51:258–266.

9. Wuttke M, Li Y, Li M, Sieber KB, Feitosa MF, Gorski M, Tin A, Wang L, Chu AY, Hoppmann A, et al. A catalog of genetic loci associated with kidney function from analyses of a million individuals. *Nat Genet*. 2019;51:957–972.

10. Doke T, Huang S, Qiu C, Sheng X, Seasock M, Liu H, Ma Z, Palmer M, Susztak K. Genome-wide association studies identify the role of caspase-9 in kidney disease. *Sci Adv*. 7:eabi8051.

11. Gillis E, Kumar AA, Luyckx I, Preuss C, Cannaerts E, Beek G van de, Wieschendorf B, Alaerts M, Bolar N, Vandeweyer G, et al. Candidate Gene Resequencing in a Large Bicuspid Aortic Valve-Associated Thoracic Aortic Aneurysm Cohort: SMAD6 as an Important Contributor. *Frontiers in physiology* [Internet]. 2017;8. Available from: https://pubmed.ncbi.nlm.nih.gov/28659821/

12. Luyckx I, MacCarrick G, Kempers M, Meester J, Geryl C, Rombouts O, Peeters N, Claes C, Boeckx N, Sakalihasan N, et al. Confirmation of the role of pathogenic SMAD6 variants in bicuspid aortic valve-related aortopathy. *European journal of human genetics : EJHG*. 2019;27:1044–1053.

13. Tan HL, Glen E, Töpf A, Hall D, O'Sullivan JJ, Sneddon L, Wren C, Avery P, Lewis RJ, ten Dijke P, et al. Nonsynonymous variants in the SMAD6 gene predispose to congenital cardiovascular malformation. *Human mutation*. 2012;33:720–727.

14. Luyckx I, Verstraeten A, Goumans M-J, Loeys B. SMAD6-deficiency in human genetic disorders. *npj Genom. Med.* 2022;7:1–11.

15. Chaffin M, Papangeli I, Simonson B, Akkad A-D, Hill MC, Arduini A, Fleming SJ, Melanson M, Hayat S, Kost-Alimova M, et al. Single-nucleus profiling of human dilated and hypertrophic cardiomyopathy. *Nature*. 2022;608:174–180.

16. Ruter DL, Liu Z, Ngo KM, X S, Marvin A, Buglak DB, Kidder EJ, Bautch VL. SMAD6 transduces endothelial cell flow responses required for blood vessel homeostasis. *Angiogenesis*. 2021;24:387–398.

17. Wylie LA, Mouillesseaux KP, Chong DC, Bautch VL. Developmental SMAD6 Loss Leads to Blood Vessel Hemorrhage and Disrupted Endothelial Cell Junctions. *Developmental biology*. 2018;442:199–199.

18. Liu C, Kraja AT, Smith JA, Brody JA, Franceschini N, Bis JC, Rice K, Morrison AC, Lu Y, Weiss S, et al. Meta-analysis identifies common and rare variants influencing blood pressure and overlapping with metabolic trait loci. *Nature genetics*. 2016;48:1162–1170.

19. Surendran P, Drenos F, Young R, Warren H, Cook JP, Manning AK, Grarup N, Sim X, Barnes DR, Witkowska K, et al. Trans-ancestry meta-analyses identify rare and common variants associated with blood pressure and hypertension. *Nature genetics*. 2016;48:1151–1161.

20. Surendran P, Feofanova EV, Lahrouchi N, Ntalla I, Karthikeyan S, Cook J, Chen L, Mifsud B, Yao C, Kraja AT, et al. Discovery of rare variants associated with blood pressure regulation through meta-analysis of 1.3 million individuals. *Nat Genet*. 2020;52:1314–1332.

21. Basu M, Sharmin M, Das A, Nair NU, Wang K, Lee JS, Chang Y-PC, Ruppin E, Hannenhalli S. Prediction and Subtyping of Hypertension from Pan-Tissue Transcriptomic and Genetic Analyses. *Genetics*. 2017;207:1121–1134.

22. Gerull B, Gramlich M, Atherton J, McNabb M, Trombitás K, Sasse-Klaassen S, Seidman JG, Seidman C, Granzier H, Labeit S, et al. Mutations of TTN, encoding the giant muscle filament titin, cause familial dilated cardiomyopathy. *Nat Genet*. 2002;30:201–204.

23. Herman DS, Lam L, Taylor MRG, Wang L, Teekakirikul P, Christodoulou D, Conner L, DePalma SR, McDonough B, Sparks E, et al. Truncations of titin causing dilated cardiomyopathy. *N Engl J Med*. 2012;366:619–628.

24. Emdin CA, Khera AV, Chaffin M, Klarin D, Natarajan P, Aragam K, Haas M, Bick A, Zekavat SM, Nomura A, et al. Analysis of predicted loss-of-function variants in UK Biobank identifies variants protective for disease. *Nature Communications* [Internet]. 2018;9. Available from: /pmc/articles/PMC5915445/

25. Chen M-H, Raffield LM, Mousas A, Sakaue S, Huffman JE, Moscati A, Trivedi B, Jiang T, Akbari P, Vuckovic D, et al. Trans-ethnic and Ancestry-Specific Blood-Cell Genetics in 746,667 Individuals from 5 Global Populations. *Cell*. 2020;182:1198-1213.e14.

26. Vuckovic D, Bao EL, Akbari P, Lareau CA, Mousas A, Jiang T, Chen M-H, Raffield LM, Tardaguila M, Huffman JE, et al. The Polygenic and Monogenic Basis of Blood Traits and Diseases. *Cell*. 2020;182:1214-1231.e11.

27. Li M, Wang A, Quek LE, Vernon S, Figtree GA, Yang J, O'Sullivan JF. Metabolites downstream of predicted loss-of-function variants inform relationship to disease. *Molecular genetics and metabolism*. 2019;128:476–482.

28. Huć T, Nowinski A, Drapala A, Konopelski P, Ufnal M. Indole and indoxyl sulfate, gut bacteria metabolites of tryptophan, change arterial blood pressure via peripheral and central mechanisms in rats. *Pharmacological Research*. 2018;130:172–179.

29. Yisireyili M, Saito S, Abudureyimu S, Adelibieke Y, Ng HY, Nishijima F, Takeshita K, Murohara T, Niwa T. Indoxyl sulfate-induced activation of (pro)renin receptor promotes cell proliferation and tissue factor expression in vascular smooth muscle cells. *PloS one* [Internet]. 2014;9. Available from: https://pubmed.ncbi.nlm.nih.gov/25343458/

30. Mansour A, Mohajeri- Tehrani MR, Qorbani M, Heshmat R, Larijani B, Hosseini S. Effect of glutamine supplementation on cardiovascular risk factors in patients with type 2 diabetes. *Nutrition*. 2015;31:119–126.

31. Zhang T, Wei H, Li M, Han W, Zhang W, Zhang X, Zhang B, Jiang Z, Li T. Risk of migraine contributed by genetic polymorphisms of ANKDD1B gene: a case-control study based on Chinese Han population. *Neurol Sci*. 2022;43:2735–2743.

32. Vujkovic M, Keaton JM, Lynch JA, Miller DR, Zhou J, Tcheandjieu C, Huffman JE, Assimes TL, Lorenz K, Zhu X, et al. Discovery of 318 new risk loci for type 2 diabetes and related vascular outcomes among 1.4 million participants in a multi-ancestry meta-analysis. *Nat Genet*. 2020;52:680–691.

33. Klimentidis YC, Arora A, Newell M, Zhou J, Ordovas JM, Renquist BJ, Wood AC. Phenotypic and Genetic Characterization of Lower LDL Cholesterol and Increased Type 2 Diabetes Risk in the UK Biobank. *Diabetes*. 2020;69:2194–2205.

34. DeBoever C, Tanigawa Y, Lindholm ME, McInnes G, Lavertu A, Ingelsson E, Chang C, Ashley EA, Bustamante CD, Daly MJ, et al. Medical relevance of protein-truncating variants across 337,205 individuals in the UK Biobank study. *Nat Commun*. 2018;9:1612.

35. Li J, Mahajan A, Tsai M-D. Ankyrin repeat: a unique motif mediating protein-protein interactions. *Biochemistry*. 2006;45:15168–15178.

36. Tan Z, Zeng H, Xu Z, Tian Q, Gao X, Zhou C, Zheng Y, Wang J, Ling G, Wang B, et al. Identification of ANKDD1B variants in an ankylosing spondylitis pedigree and a sporadic patient. *BMC Medical Genetics* [Internet]. 2018;19. Available from: /pmc/articles/PMC6034262/

37. Yang Y, Zhao H, Boomsma DI, Ligthart L, Belin AC, Smith GD, Esko T, Freilinger TM, Hansen TF, Ikram MA, et al. Molecular genetic overlap between migraine and major depressive disorder. *Eur J Hum Genet*. 2018;26:1202–1216.

38. Guo Y, Rist PM, Daghlas I, Giulianini F, Kurth T, Chasman DI. A genome-wide cross-phenotype meta-analysis of the association of blood pressure with migraine. *Nat Commun*. 2020;11:3368.

39. Ryu CS, Sakong JH, Ahn EH, Kim JO, Ko D, Kim JH, Lee WS, Kim NK. Association study of the three functional polymorphisms (TAS2R46G>A, OR4C16G>A, and OR4X1A>T) with recurrent pregnancy loss. *Genes Genom*. 2019;41:61–70.

40. Takabatake N, Toriyama S, Takeishi Y, Shibata Y, Konta T, Inoue S, Abe S, Igarashi A, Tokairin Y, Ishii M, et al. A nonfunctioning single nucleotide polymorphism in olfactory receptor gene family is associated with the forced expiratory volume in the first second/the forced vital capacity values of pulmonary function test in a Japanese population. *Biochemical and Biophysical Research Communications*. 2007;364:662–667.

41. Chen Z, Zhao H, Fu N, Chen L. The diversified function and potential therapy of ectopic olfactory receptors in non-olfactory tissues. *J Cell Physiol*. 2018;233:2104–2115.
